# Assessing the impacts of timing on the health benefits, cost-effectiveness and relative affordability of COVID-19 vaccination programmes in 27 African Countries

**DOI:** 10.1101/2022.05.09.22274846

**Authors:** Yang Liu, Carl AB Pearson, Andrés Madriz Montero, Sergio Torres-Rueda, Elias Asfaw, Benjamin Uzochukwu, Tom Drake, Eleanor Bergren, Rosalind M Eggo, Francis Ruiz, Nicaise Ndembi, Justice Nonvignon, Mark Jit, Anna Vassall

## Abstract

**Background:** The COVID-19 vaccine supply shortage in 2021 constrained rollout efforts in Africa while populations experienced waves of epidemics. As supply picks up, a key question becomes if vaccination remains an impactful and cost-effective strategy given changes in the timing of implementation.

**Methods:** We assessed the impact of timing using an epidemiological and economic model. We fitted our mathematical epidemiological model to reported COVID-19 deaths in 27 African countries to estimate the existing immunity (resulting from infection) before substantial vaccine rollout. We then projected health outcomes for different programme start dates (2021-01-01 to 2021-12-01, n = 12) and roll-out rates (slow, medium, fast; 275, 826, and 2066 doses/ million population-day, respectively) for viral vector and mRNA vaccines. Rollout rates used were derived from observed uptake trajectories. We collected data on vaccine delivery costs by country income group. Lastly, we calculated incremental cost-effectiveness ratios and relative affordability.

**Findings:** Vaccination programmes with early start dates incur the most health benefits and are most cost-effective. While incurring the most health benefits, fast vaccine roll-outs are not always the most cost-effective. At a willingness-to-pay threshold of 0.5xGDP per capita, vaccine programmes starting in August 2021 using mRNA and viral vector vaccines were cost-effective in 6-10 and 17-18 of 27 countries, respectively.

**Interpretation:** African countries with large proportions of their populations unvaccinated by late 2021 may find vaccination programmes less cost-effective than they could have been earlier in 2021. Lower vaccine purchasing costs and/or the emergence of new variants may improve cost-effectiveness.

**Funding:** Bill and Melinda Gates Foundation, World Health Organization, National Institute of Health Research (UK), Health Data Research (UK)

## Introduction

Since COVID-19 vaccines were first authorised in late 2020, countries with bilateral deals with vaccine manufacturers mostly achieved considerable coverage in a short period, enormously reducing disease and economic burdens. However, accruing COVID-19 vaccine coverage in many African countries has been challenging as they have relied on the COVAX initiative to procure vaccines. Based on recent data on roll-out efforts, it will be a substantial challenge for the region to achieve the 70% coverage target by mid-2022 as set out by the World Health Organization (WHO) and the Africa Centres for Disease Control and Prevention (Africa CDC).(1–5)

Until late 2021, delays in COVID-19 vaccination were largely driven by vaccine supply constraints. However, the supply available through COVAX has expanded substantially since early 2022.(3) By this time, however, many African Union (AU) member states had already experienced several epidemic waves involving multiple variants of concern (VOCs) at considerable health and economic costs to their populations.(1,6)

A key question that AU member states now face is whether rolling-out COVID-19 vaccines remains an impactful strategy representing good value for money, having missed key windows of opportunities from early-to mid-2021. Continuing vaccination raises issues in respect of both cost-effectiveness and affordability in the region since the investment needed may incur substantial health opportunity costs for other health services. (7) Both costs and effects associated with vaccination programmes may vary by the timing of implementation.

This study aims to inform future decisions about vaccine roll-out and investment by retrospectively examining the impacts of implementation timing on COVID-19 disease burden using a combined mathematical and economic modelling approach. This approach allowed us to factor in the potentially high seroprevalence of the region, the emergence of multiple variants of concern (VOCs), and population characteristics key to the transmission of SARS-CoV-2 (e.g. population age structure, contacts) and to the economic evaluation of vaccination programmes (Gross Domestic Product per capita (GDPpc), general public health expenditures). (8–10) We estimated the health impacts, cost-effectiveness, and relative affordability by vaccine types across the AU while highlighting the main drivers of differences. We estimated both cost-effectiveness and relative affordability as we are concerned about the large non-marginal impact not captured by the cost-effectiveness related decision-making threshold.

This is the first multi-country regional analysis of COVID-19 vaccine strategies for Africa that links epidemiological models and economic evaluation. The lessons learned could inform national decision-makers on future vaccine roll-out decisions in Africa and globally, particularly if reformulated vaccines become available in response to future VOCs. To our knowledge, this is also the first study to examine the impact of implementation timing while appraising COVID-19 vaccine policies, with findings that reflect the heterogeneity of the African region.

## Methods

We estimated the prevalence of infection-induced protection against COVID-19 within populations by fitting the dynamic transmission model to country-level daily reported COVID-19 deaths. Cumulative health outcomes and disability-adjusted life years (DALYs) averted associated with different vaccine roll-out scenarios were then calculated. Combining health outcomes with data on the costs of vaccine delivery and COVID-19 related health service, we then estimated the overall cost-effectiveness and relative affordability of each vaccine roll-out scenario from a health sector perspective relying on the between-country heterogeneity as an important source of uncertainty. This section provides essential information on these procedures and further details are presented in the Supplemental Materials.

### Characterising vaccine roll-out scenarios

By the time most AU members achieved 1% COVID-19 vaccine coverage (i.e. after August 2021), countries like the United Kingdom had vaccinated 74.6% (as of 31 August 2021) of its population.(1) By February 2022, vaccine coverage in over 35% of AU members was still below 10% (Figure 1a). (1) We explored possible alternative vaccine rollout trajectories that could have taken place in 2021 by varying two parameters: vaccination programme start date and vaccine roll-out rates. The programme start date was defined as the date at which the first person in a country was vaccinated and was varied between 01 January 2021 to 01 December 2021 in one-month increments. Roll-out rate was defined as the number of people vaccinated per million population-day and was varied between three levels derived based on the median observed vaccine roll-out rates of the region by tertile (275, 826, and 2066 doses/ million population-day, hereafter labelled as “slow”, “medium”, and “fast”, respectively) (methods on derivation are presented in Supplemental Methods p32). The vaccine roll-out scenarios combining these two parameters (n=36) are visualised in Figure 1b. We capped the maximum population-level vaccine coverage at 70%, consistent with the target set by the WHO and the Africa CDC. (2,5) Older adults (60+ years of age) were prioritised in the vaccine roll-out process.

**Figure 1.**
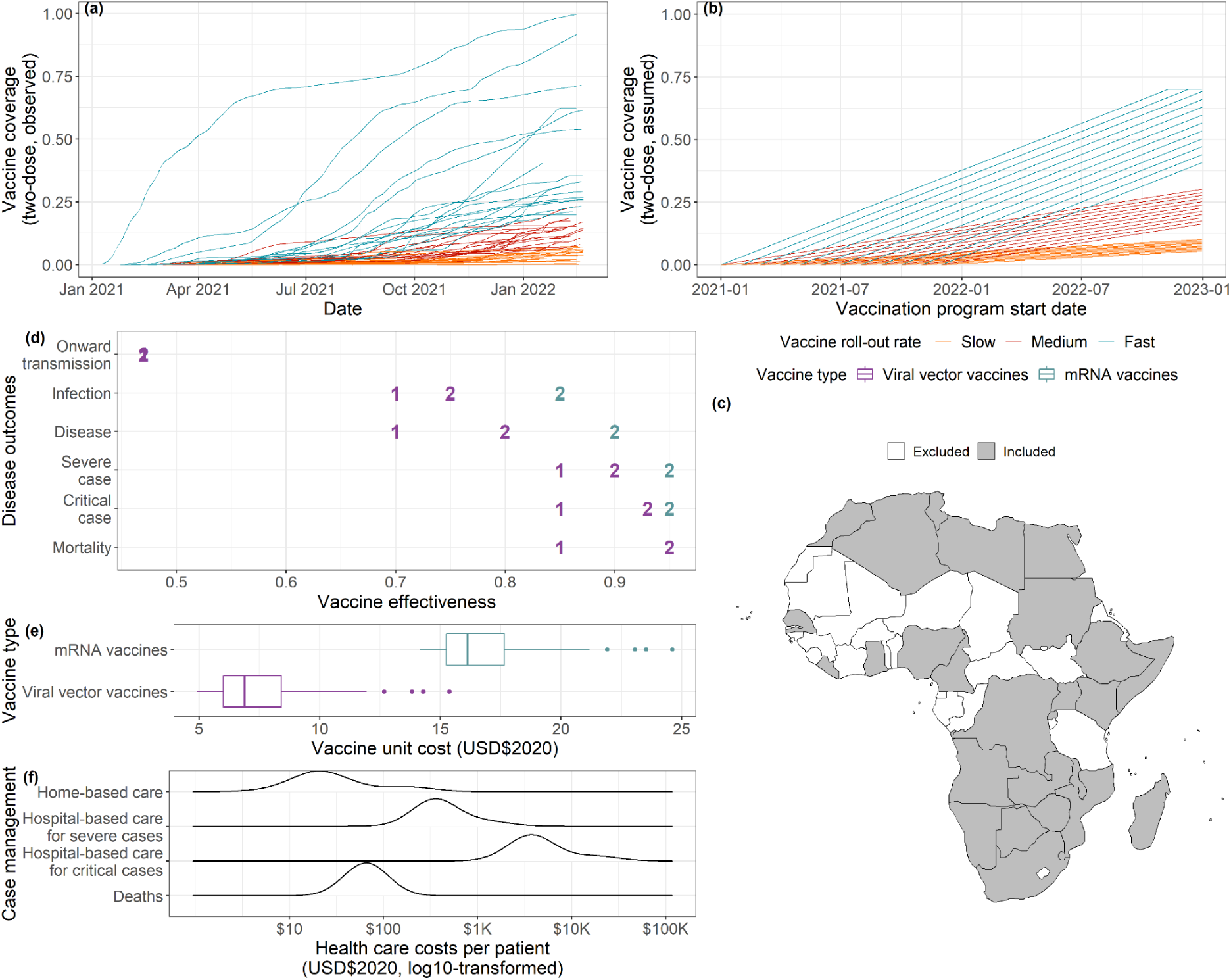
Model parameterisation and vaccine roll-out scenarios set-up. (a) Vaccine uptake observed among African Union members. Each line represents a country, and its colour indicates the roll-out rate as categorised using the methods described in the Supplemental Methods XXXX. (b) Vaccine uptake corresponding to each of the 36 vaccine roll-out scenarios (combinations of vaccination program start dates and vaccine roll-out rates, n = 36) and the population level vaccine coverage levels they realised. (c) African Union members included in the analysis. (d) Base case vaccine effectiveness estimates. Dose numbers are represented by “1” and “2”. Colours indicate vaccine type. Specific values and justifications are presented in Supplemental Table S7. (e) Distributions of vaccine unit costs among Africa Union members by vaccine type based on data collected for this study. Colours indicate vaccine type. This is based on “medium” roll-out scenarios and an 18 month-long vaccination campaign duration. (f) Distributions of health care unit costs among African Union members for case management at home and in hospitals based on values presented in Torres-Rueda et al. (11)

### The transmission model

The epidemic model used in this study was an adaptation of CovidM, an age-specific dynamic transmission model of COVID-19 which has been previously used to explore the impact of different vaccination strategies in the UK, European countries, and Pakistan (Supplemental Figure S1, Supplemental Table S1-S4, Supplemental Methods p33, p35).(12–14) We estimated the following parameters by fitting this model to country-level daily reported COVID-19 deaths: (1) the basic reproduction numbers (R0); (2) infection introduction dates; (3) COVID-19 death reporting rate; and (4) VOC introduction dates (Supplemental Table S5-S6, Supplemental Methods p34). Among the 55 AU members, we identified 27 members with sufficient data for model fitting (Figure 1c). The remaining 28 AU members were excluded from further analysis due to either data sparsity (i.e. <= 10 deaths/ day throughout the fitting period, n = 26) or potential reporting artefacts (i.e. with single day accounting for > 5% of the cumulative COVID-19 deaths since early 2020, n = 2).

We explored vaccine rollout scenarios with either mRNA vaccines (with characteristics similar to those of the Pfizer-BioNTech COVID-19 vaccines) or viral vector vaccines (with characteristics similar to those of Oxford/AstraZeneca vaccines). Five types of vaccine effect mechanisms were incorporated into the model: infection-, disease-, severe case-, critical case-, mortality-reducing and onward transmission-blocking. The specific vaccine effectiveness (VE) values are presented in Figure 1d, and the justification behind them in Supplemental Table S7. We tested an additional set of VE values (see Supplemental Table S8) using the lower bounds of the uncertainty around the VE estimates in existing literature as a sensitivity analysis.

We projected four health outcomes under these vaccine roll-out scenarios: (1) symptomatic infections; (2) severe cases that require hospitalisation; (3) critical cases that require intensive care unit (ICU) admission; and (4) deaths (see Supplemental Methods p36). Regardless of the roll-out scenario, all outcomes were aggregated from 01 January 2021 to 31 December 2022. We kept the time horizon relatively short due to the uncertainty around future VoCs. However, we extended it to 30 June 2023 as a sensitivity analysis as vaccination programs that start late may require time to present impacts.

### Calculating DALYs

We used projected health outcomes to calculate DALYs incurred, which capture disease burden in terms of both mortality and morbidity. Mortality is measured as Years of Life Lost (YLLs) per COVID-19 death by combining model projections of age-specific COVID-19 deaths with remaining life expectancy in the absence of COVID-19 using country-specific life tables.(9) In a sensitivity analysis, we assumed that individuals who die of COVID-19 have risk factors that may have predisposed them to die earlier even if they had not acquired COVID-19. Under this assumption, we recalculated the YLLs using the life expectancy with a 50% increase in the age-specific mortality (i.e. Standardised Mortality Ratio = 1.5). Morbidity is measured as Years Lived with Disability (YLDs) among cases (i.e. symptomatic infections), severe cases, critical cases and those who experience long-term health effects (“Long-COVID”).

We discounted YLLs over remaining life expectancy and additionally discounted DALYs according to the year in which COVID-19 health outcomes occurred. In line with the WHO guidelines on the economic evaluation of immunisation programmes, we used an annual discount rate of 3% in the base case analysis and used 0% discounting as a sensitivity analysis. (15) More information on calculating DALYs can be found in Supplemental Methods p38, p40, and p42.

### Estimating the vaccine delivery and health service costs

We collected data on country-specific unit costs per vaccine delivered by vaccine type from a healthcare provider perspective in three countries (Ethiopia (low-income country), Nigeria (lower-middle-income country), and South Africa (upper-middle-income country)) using a normative ingredient-based approach (Supplemental Methods p43).(16,17) The unit cost includes vaccine purchasing costs and related components and activities involved in the planning and delivery of the vaccine (e.g. planning and coordination, cold chain, transportation and waste disposal as defined in the COVAX Working Group on delivery costs. (16). Costs were then validated by country-level experts knowledgeable of the immunisation effort. In the case of Ethiopia and Nigeria, this local validation was part of the health technology assessment process used to support decision making around COVID-19 vaccinations.

Based on raw data collected, the unit cost of delivering mRNA vaccines is approximately two times of delivering viral vector vaccines (Figure 1e). The unit cost differences by vaccine type are driven by vaccine purchasing costs (see itemized costs by country, activity, and component in Supplementary Methods 43, Supplemental Tables S9-S10). We extrapolated these vaccine unit costs for other countries (see Supplementary Methods p52) and other vaccination programme setups (i.e. programme duration and daily vaccine roll-out rate, see Supplementary Methods p54, Table S11).

For COVID-19 related health sector costs, we used previously published country-specific estimates (for home-based care, hospital-based care for severe cases, hospital-based care for critical cases, and management of fatal cases) and lengths of hospital stay (Figure 1f). (11) All costs were converted to US$2020. Similarly to DALYs, costs were discounted by 3% in line with WHO guidelines (Supplemental Methods p42).

### Measuring cost-effectiveness and relative affordability

We estimated incremental cost-effectiveness ratios (ICERs) from a healthcare perspective for different vaccination scenarios compared to a no vaccination scenario by dividing the differences in total costs between alternatives (including vaccine delivery and COVID-19 health service costs) by the differences in DALYs incurred.

We assess cost-effectiveness relative to an opportunity cost threshold that varies across countries broadly related to a country’s GDP per capita (GDPpc)(18,19). To explore patterns of cost-effectiveness across countries, we divided the ICERs by GDP per capita (GDPpc) to quantify the relative value for money of different vaccine roll-out scenarios, generating what we term ‘normalised’ ICERs (nICERs). We used an nICER of 0.5 as the base case decision threshold (nICER estimates of 0.5 or less were considered cost-effective), and we used 0.3 and 1 as alternative thresholds in the sensitivity analysis.

In this study, we defined the relative affordability of a vaccine programme in a given year as the annual incremental cost of the programme (i.e. costs of rolling-out vaccinations minus health service expenditure offset) divided by annual government general health expenditure.(20)

All analysis has been done in R (4.1.0). Code used and intermediate results are publicly available at https://github.com/yangclaraliu/covid_vac_africa.

## Results

Using the fitted models (see Supplemental Figure S2), we projected the cumulative cases, severe cases, critical cases and deaths occurring between 01 January 2021 and 31 December 2022 under different vaccination scenarios. We present the health impacts of viral vector vaccines on cases and deaths in Figure 2. Results for mRNA vaccines and for additional health outcomes can be found in Supplemental Figure S3-S5. Our discussion centres around relative reduction in disease burden instead of absolute differences to allow the comparison across countries with varying population sizes.

**Figure 2.**
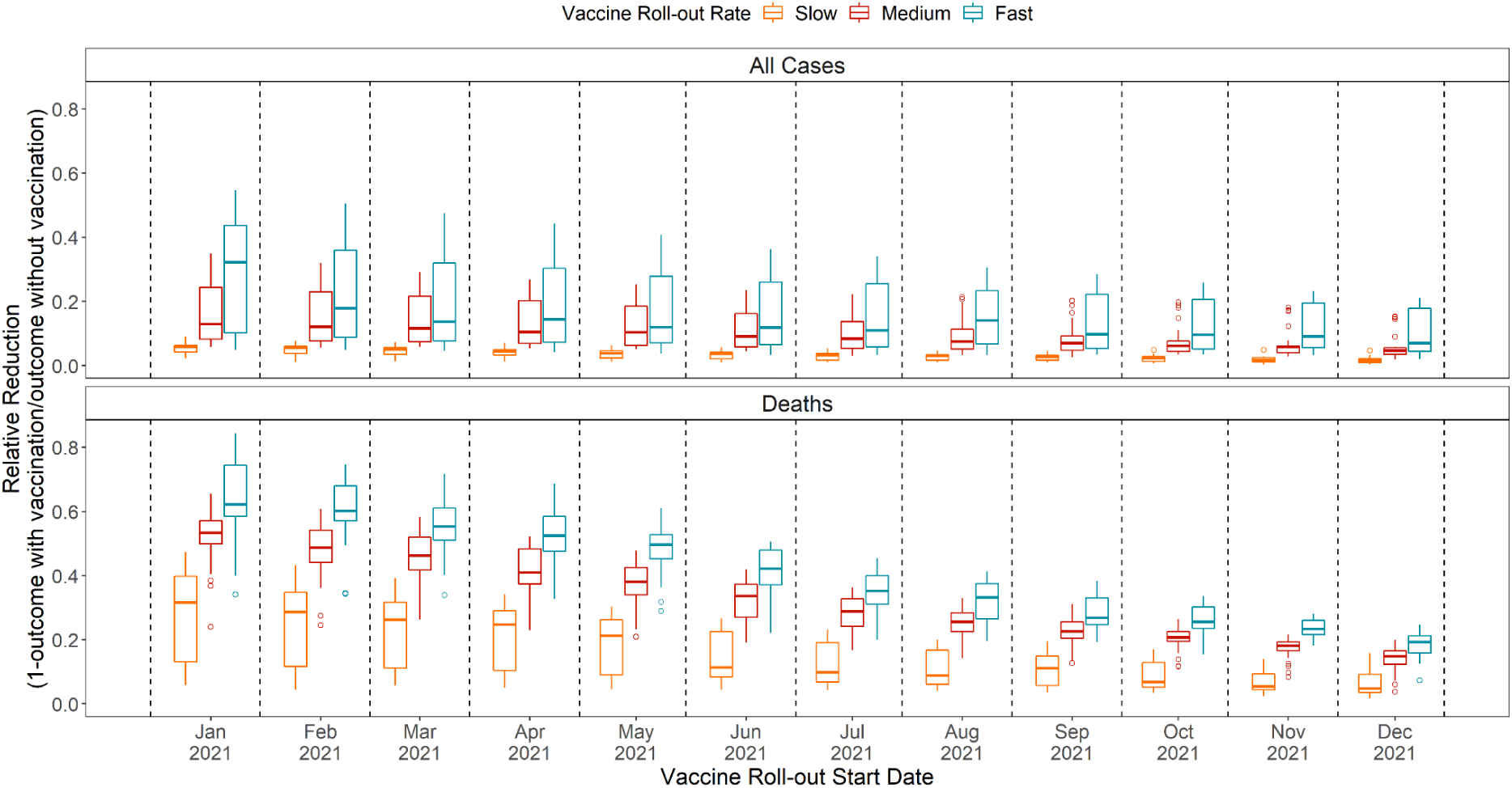
Health Outcomes Associated with Different Vaccine Roll-out Scenarios for 27 African Union Members using viral vector vaccines. Relative reduction in health burden as a result of different vaccine roll-out scenarios (i.e. combinations of vaccination program start dates and vaccine roll-out rates) for 27 AU members. Relative reduction is defined as 1 - outcome with vaccination/outcome without vaccination. Greater relative reductions indicate more effective vaccine roll-out scenarios, and vice versa. Results for additional health outcomes (severe and critical cases) and vaccine types can be found in Supplemental Figures S3-S5.

For a given vaccination program start date, faster roll-out rates are associated with a substantially greater disease burden averted compared to the no vaccination baseline. For example, compared to no vaccination, vaccination programmes starting in August 2021 (when more than half of AU members with data (n = 53) reached 1% population-level vaccine coverage) could have reduced deaths by an average of 11.06% [median = 8.80%, IQR: 5.87% - 16.20%; n = 27 (countries)] if vaccination had been rolled out under the slow scenario, 24.19% [24.43%, 21.78% - 27.18%] under the medium scenario, and 31.29% [32.57%, 25.72% - 36.68%] under the fast scenario.

For a given vaccine roll-out rate, earlier roll-out start dates are associated with greater disease burden averted compared to the no vaccination baseline. For example, using the medium roll-out rate, starting in January could have reduced deaths by a mean of 50.30% [51.29%, 48.97% - 54.95%]. However, starting the same programme in August 2021 could only have reduced deaths by a mean of 24.19% [24.32%, 21.78% - 27.18%]. The relative differences between different roll-out rates were greater for programmes with earlier start dates.

Given the same start date, faster programmes are associated with higher nICERs (i.e. are less cost-effective); using a given vaccine roll-out rate, later programmes are associated with higher nICERs (Figure 3). Additionally, we found that few countries would find programmes using mRNA vaccines to be cost-effective. There were 10, 8, and 6 (of 27 countries) countries that would find vaccination programmes starting from August 2021 to be cost-effective while implementing slow, medium and fast roll-out rates using mRNA vaccines. Using viral vector vaccines, these values increased to 18, 18 and 17 countries (of 27 countries).

**Figure 3.**
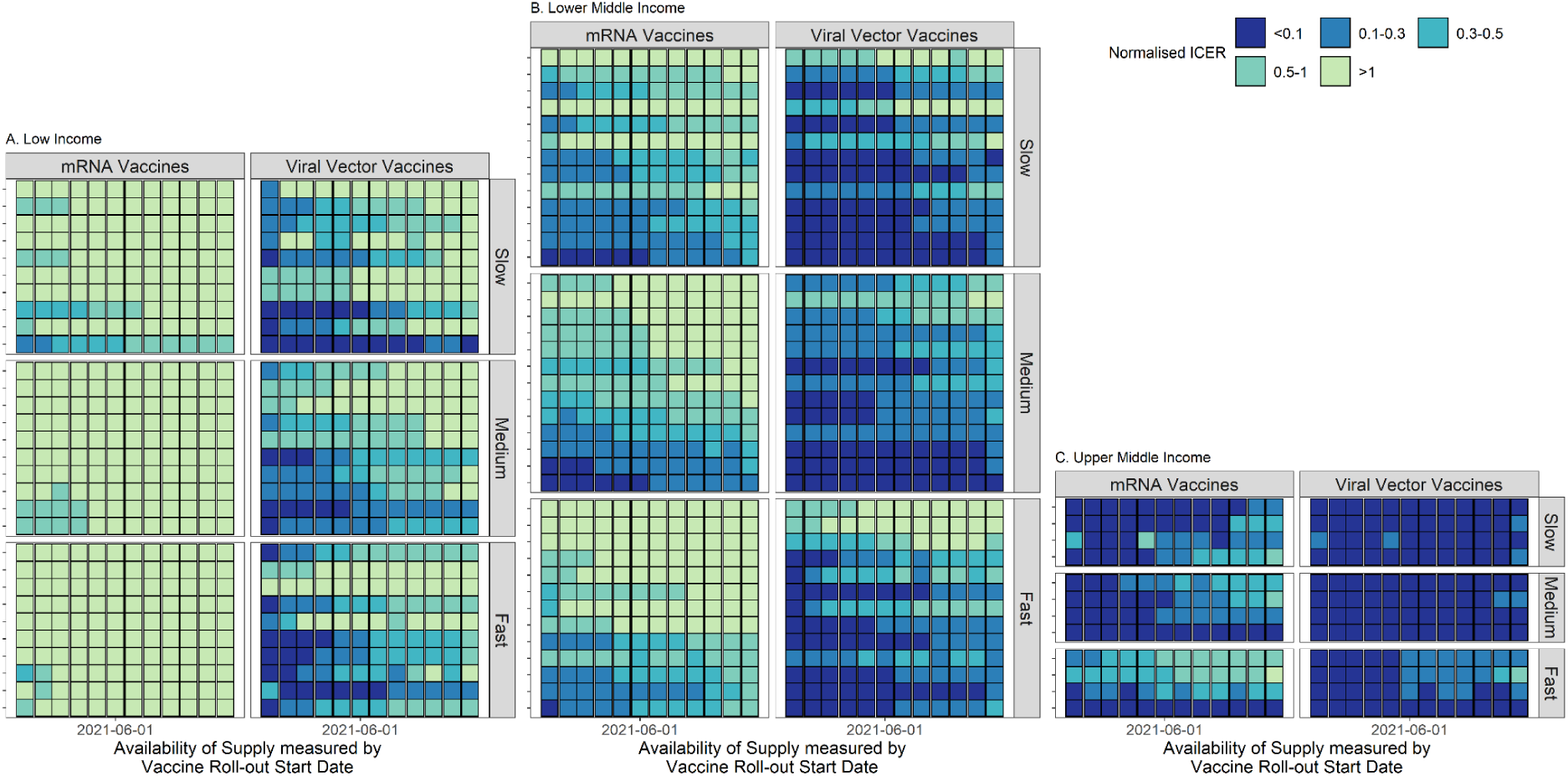
Normalised ICERs (nICERs) of vaccine roll-out scenarios compared to the no vaccination scenario (01 Jan 2021 - 31 Dec 2022). A. low-income countries; B. lower-middle-income countries; and C. upper-middle-income countries. Normalised ICER is visualised here instead of ICER as we are interested in the variations in policy evaluation outcomes after factoring in decision-making criteria (i.e. GDPpc-based thresholds). Each y-axis tick represents a country; the y-axis is arranged based on nICER values.

Comparison across income groups shows that the lowest nICERs are observed in upper-middle-income countries, followed by lower-middle and low-income countries (Figure 3). Both mRNA and viral vector vaccine-based programmes may be cost-effective in upper-middle-income countries in most roll-out scenarios (Figure 3C). However, only viral vector vaccines and only early roll-out scenarios may be cost-effective in low-income countries (Figure 3A).

Our sensitivity analyses around the time horizon show that drawing the line at 31 Dec 2022 may lead to higher estimates in the proportions of countries with cost-effective vaccination scenarios given a later start date (Figure 4a). The decreasing trends observed in Figure 2 as vaccination programmes delay may be steeper, and the contrast between early and late starting programs (in Figure 3A-B) more evident as the time horizon extends. Using VE estimates corresponding to the lower bounds of confidence ranges found in literature only affected our findings for viral vector vaccines (Figure 4b). This reflects a more drastic drop in the VE used for viral vector vaccines than mRNA vaccines (Supplemental Table S7-S8). Proportions of countries that would consider a given vaccine roll-out scenario cost-effective are also strongly affected by the decision making criteria (Figure 4c) and economic evaluation scenarios (Figure 4d).

**Figure 4.**
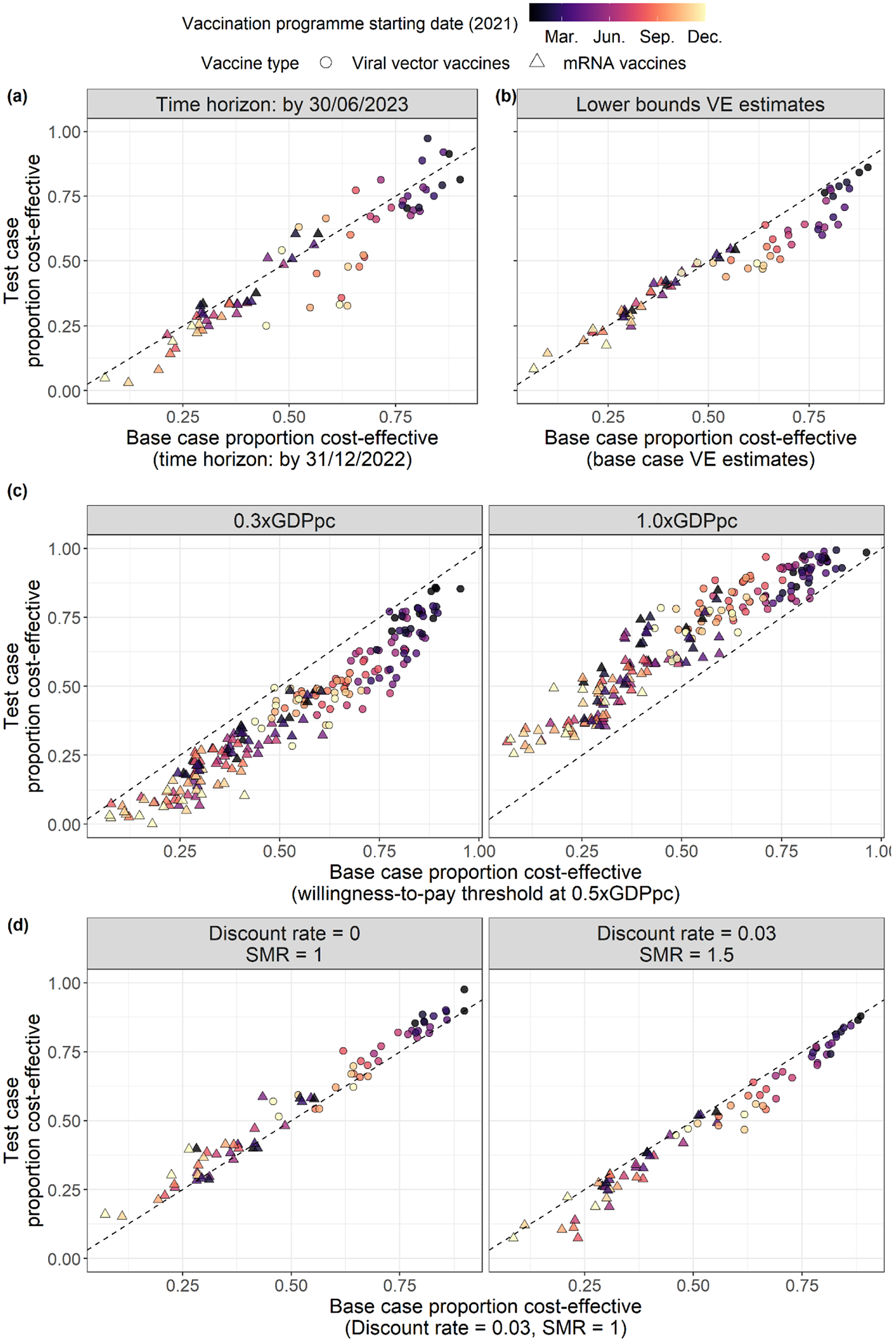
Results from sensitivity analyses. We present results from sensitivity analyses around (a) time horizon; (b) vaccine effectiveness; (c) decision making criteria (i.e. GDPpc-based thresholds); and (d) economic evaluation scenarios while calculating DALYs. Point shapes indicate vaccine type, and colours indicate vaccination programme start date. For each sensitivity analysis, we include the vaccine roll-out rate dimension in the Supplemental Figures S6-S9. In this figure, each point represents a given vaccine type, vaccination programme start date and vaccine roll-out rate (n = 2*12*3 points per panel). SMR: standardized mortality ratio.

The median relative affordability estimates for vaccination programmes involving mRNA vaccines implemented with slow, medium, and fast roll-out rates are 3.85% [IQR: 1.26%-10.24%], 12.42%[3.43%-29.84%], and 26.13%[7.82%-61.61%] of the domestic general government health expenditure, respectively; and for viral vector vaccines 1.09%[0.18%-3.02%], 3.24%[0.61%-6.61%], and 5.28%[0.70%-10.98%] of the domestic general government health expenditure, respectively (Figure 5). While cost-effectiveness and relative affordability seem to correlate on the regional level, there are two countries where some cost-effectiveness vaccine roll-out scenarios (i.e. nICER <0.5) may be associated with relative affordability above 20%. Using cost-effectiveness as the sole programme evaluation criteria may risk large opportunity costs for other public health and health service investment issues (e.g. routine childhood immunisation).

**Figure 5.**
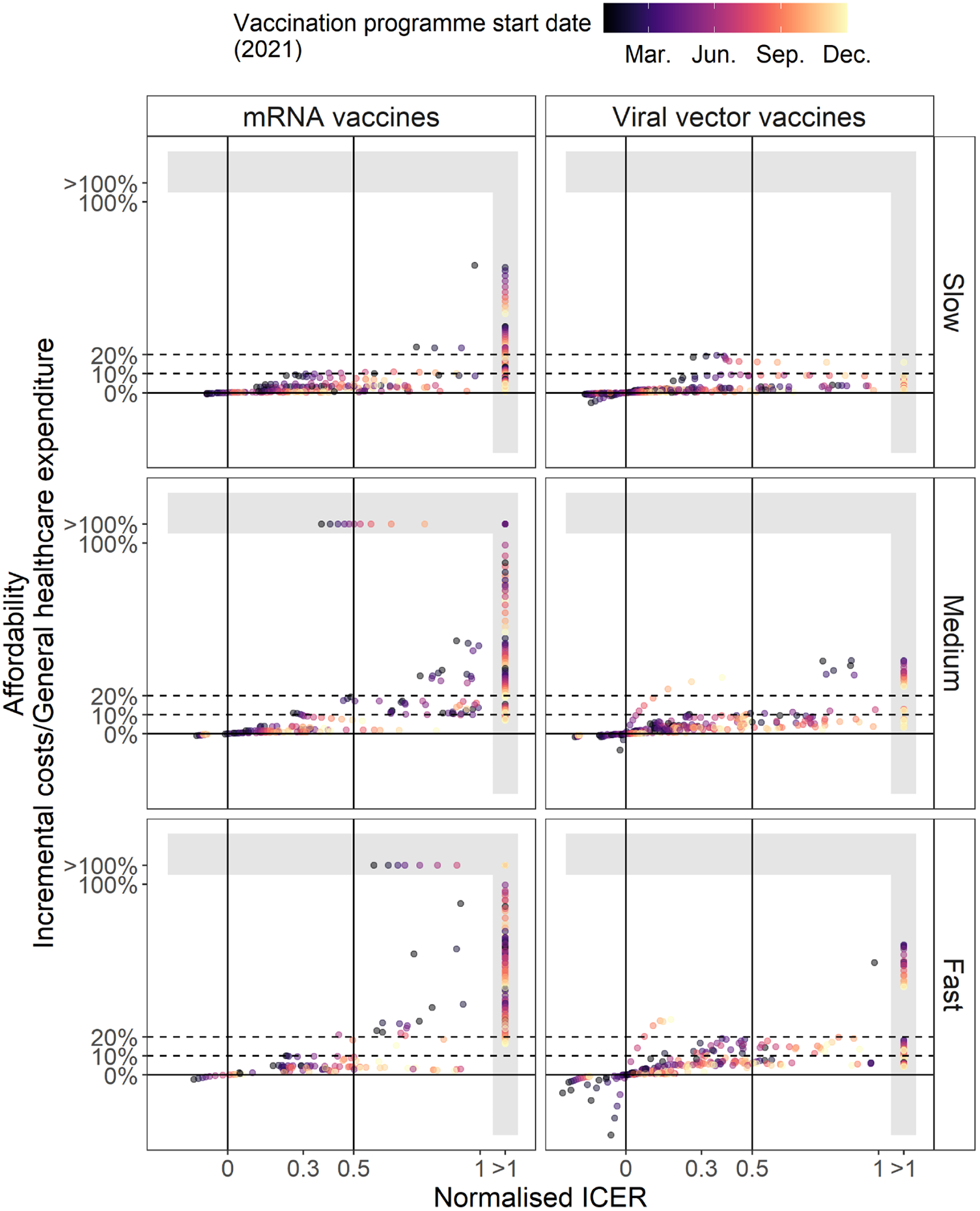
Normalised ICERs and relative affordability of COVID-19 vaccine roll-out scenarios. There are some large values in both relative affordability (max = 321.31%) and nICER (max = 15.26). We collapse relative affordability greater than 100% and nICER greater than 1 as discrete categories to visualise their relationships at ranges most relevant for vaccine policy. Negative relative affordability indicates cost-saving.

## Discussion

Using a combined epidemiological and economic modelling approach, we explored how the timing of implementation (i.e. vaccination programme start date and vaccine roll-out rates) affects vaccination programmes’ health benefits, cost-effectiveness, and relative affordability among African countries. We found that vaccination programmes with earlier start dates lead to greater health benefits and are more likely cost-effective compared to the no vaccination scenario. Fast vaccination roll-out leads to the greatest health benefits (compared to medium and slow scenarios) but is not always the most cost-effective. This is likely due to diminishing returns on investment as fast scenarios quickly move beyond the high-risk and thus prioritised population (defined as 60+ years of age in this study).

In some low-income and lower-middle-income countries, late start and slow vaccine roll-out scenarios may not be cost-effective (using a 0.5xGDPpc threshold) and have poorer value for money due to the epidemic moving faster than vaccination in terms of creating immunity. There were missed opportunities in early-mid 2021 for COVID-19 vaccination to save more lives in Africa. In countries with large proportions of their populations unvaccinated by the end of 2021, vaccinating people is now (in mid-2022) less cost-effective than in early-mid 2021, given the immune-evading nature of the Omicron variant and the high seroprevalence in the region. However, two factors may significantly improve the overall cost-effectiveness of vaccination programmes. First, our results are based on the known immune dynamics of SARS-CoV-2 and the performance of current COVID-19 vaccines. New vaccines with better effectiveness against emerging VOCs could improve cost-effectiveness. Emerging VOCs with the protection advantage of vaccine-induced immunity over infection-induced immunity may have the same effect. Second, our results have shown that vaccine roll-out scenarios are much more likely to be cost-effective when using viral vector vaccines than when using mRNA vaccines. The similar VE estimates (especially against serious health outcomes, see Figure 1d and Supplemental Table S7) and yet drastically different vaccine unit costs are likely to explain this outcome. The main driver of vaccine unit costs is vaccine price. If AU members could purchase COVID-19 vaccines at significantly lower prices, or receive more subsidised or free vaccines, then overall cost-effectiveness across scenarios and the region from a domestic opportunity cost perspective would greatly improve. However, if the funds used by development partners to provide subsidised or free vaccines replace funding for other essential health services in Africa, opportunity costs may still be too high.

We found that the most cost-effective vaccine roll-out scenarios were also relatively affordable. However, in a few countries-and-vaccine-roll-out-scenario combinations appeared cost-effective but relatively unaffordable. The implication is that the investment in COVID-19 vaccination may risk opportunity costs for other public health and health service investment issues. We have likely underestimated the affordability issue in this study as we have not explicitly accounted for the negative impacts of the pandemic on the local health systems and essential health services (e.g. the impact of lockdown). Temporary external funding to support the local public health expenditure may help make COVID-19 vaccination programmes more affordable.

Our results are sensitive to decision-making criteria and metrics. The decision-making criteria we used in this study are not based on local decision rules but empirical assessment of opportunity cost. We used the 0.5xGDPpc threshold for cost-effectiveness and did not use a previously commonly-cited threshold of 1xGDPpc, as this has been shown to significantly underestimate the opportunity costs in all lower-income countries and most middle-income countries.(18) However, opportunity costs are likely to be higher for non-marginal interventions (e.g. those with a large budget impact). Therefore even with a 0.5xGDPpc, we may overestimate cost-effectiveness. Our sensitivity analysis demonstrated that if decision-makers decide to use a different threshold, the overall cost-effectiveness across the region would be substantially altered. We also presented the paired cost-effectiveness and relative affordability results to inform vaccination programmes considering multiple metrics.(21) However, each country’s decision should rest on its own assessment and appraisal of both criteria and others, including equity consideration.

This is the first study to examine the importance of timing in the cost-effectiveness of COVID-19 vaccination programmes. Timing has already been shown to strongly influence health benefits of vaccination programmes, and incorporating it into economic evaluation would lead to more accurate and realistic cost-effectiveness assessments.(22) We fit a mathematical model with a range of plausible vaccine effect mechanisms to the COVID-19 epidemic histories in 27 AU members. Most existing modelling studies on the economic evaluation of COVID-19 vaccination strategies have either focused on a few high-income countries or used hypothetical epidemic histories that are not based on data from any country, which hinders the interpretability and generalisability of their results. This study focused the discussion on the regional level and provided insights into optimal vaccine roll-out efforts specifically for countries with substantial resource constraints. Besides using the observed epidemic histories, we also integrated multiple data sources describing country-specific characteristics crucial to SARS-CoV-2 transmission and vaccine evaluation, including age structure, social contacts, life expectancy, GDPpc, vaccine unit costs, and health care service costs.

Our study has several limitations. First, our epidemic model may not have captured the full complexity of the immune dynamics against SARS-CoV-2, which involves both vaccination and infections. For example, our model assumed that vaccinated individuals who had prior infection histories are completely protected against SARS-CoV-2. This design is intended to make sure that most individuals within the model receive a maximum of two doses. However, we cannot capture a small number of breakthrough infections that may still happen. However, these tend to be mild cases and thus should not affect our results substantially. (23) Second, the vaccine delivery and healthcare costs used for most countries in our study were extrapolated based on a small number of countries of different income groups, which may introduce bias. More accurate country-specific cost data should be incorporated. Third, for calculating YLLs, there was a lack of empirical evidence on the baseline life expectancy for COVID-19 deaths compared to that of the general population, although we explored different assumptions in the sensitivity analysis. Further research on COVID-19 death risk profiles may provide further insights into their differences. Fourth, our costs do not account for increased prices of scarce resources that may be associated with the pace of the roll-out, which would increase opportunity costs. Moreover, we do not use observed costs of different vaccination programmes, as this data is not yet available. Finally, the cost-effectiveness analysis we presented is from a national perspective and does not capture the externalities of COVID-19 transmission in other countries (e.g. infection spillover), which may be an important consideration for funders outside the region.

## Conclusion

We assessed the impact of timing on the health benefits, cost-effectiveness, and relative affordability of COVID-19 vaccination programmes in 27 African countries. We found that earlier vaccination programmes led to greater health benefits and were more cost-effective. Although fast vaccination programmes were associated with the greatest health benefits, they were not always the most cost-effective as they involved vaccinating larger proportions of individuals with low risks for serious COVID-19 outcomes. Lower vaccine purchasing costs, improved vaccine effectiveness, and temporary external funding supporting general public health practices (e.g. childhood routine vaccination) may improve the overall cost-effectiveness and affordability of COVID-19 vaccination programmes.

## Data Availability

We used publicly available aggregate data in this study, cited in the reference list or the Supplemental Material. The CovidM modelling framework has been published previously and is available on the CMMID COVID-19 GitHub page. All code used and country-specific intermediate results are publicly available at https://github.com/yangclaraliu/covid_vac_africa. A CHEERS checklist (2022 version) is presented in Supplemental Table S12.

## Author contributions

YL, NN, JN, MJ, AV, RME and TD conceived the idea. YL, JN, MJ, and AV designed the study. YL, AMM, ST-R, BSC, EB collected the data. YL, CABP, AMM, and ST-R analysed the data, built the models, and conducted the analysis. YL conducted the literature search and visualised the data and results. YL wrote the manuscript. All co-authors have contributed to interpreting the results, reviewing and providing feedback to the manuscript.

## Acknowledgement

The authors acknowledge the contribution of Dr Nicholas G. Davies and Dr Rosanna C. Barnard for their contribution to the development of the general CovidM framework. The authors also thank Dr Simon R. Procter for his feedback on this manuscript.

