## Supplemental Material for "Assessing the impacts of timing on the health benefits, cost-effectiveness and relative affordability of COVID-19 vaccination programmes in 27 African Countries"

### Table of Content

**Supplemental Figures** **4**

Figure S1. Diagram of Transmission Model Structure 4

Figure S2. Performance of model fitting process 5

Figure S3. Health Outcomes Associated with Different Vaccine Roll-out Scenarios for 27 African Union Members (viral vector vaccines, severe and critical cases) 6

Figure S4. Health Outcomes Associated with Different Vaccine Roll-out Scenarios for 27 African Union Members (mRNA vaccines, cases and deaths) 7

Figure S5. Health Outcomes Associated with Different Vaccine Roll-out Scenarios for 27 African Union Members (mRNA vaccines, severe and critical cases) 8

Figure S6. Results from sensitivity analyses by vaccine rollout rates: time horizon of outcome aggregation 8

Figure S7. Results from sensitivity analyses by vaccine rollout rates: vaccine effectiveness estimates 10

Figure S8. Results from sensitivity analyses by vaccine rollout rates: willingness-to-pay/ decision-making criteria 11

Figure S9. Results from sensitivity analyses by vaccine rollout rates: economic evaluation parameter combinations 13

**Supplemental Tables** **15**

Table S1. Model Equations 15

Table S2. Epidemic and Healthcare Process Parameters* 18

Table S3. Additional Data Sources* 19

Table S4. Other vaccine and vaccination program characteristics 20

Table S5. Variants of Concern Introduction 21

Table S6. List of countries with fitted models 22

Table S7. Vaccine Efficacy (Base case) 24

Table S8. Vaccine Efficacy (lower limit, used for sensitivity analysis) 25

Table S9. Itemized cost per dose per activity for base countries - viral vector / AZ-like vaccine - USD$ 2021 26

Table S10. Itemized Cost per dose per activity for base countries - mRNA / Pfizer-like vaccine - USD$ 2021 27

Table S11. Cost per dose for countries with fitted models - USD$ 2021 28

Table S12. CHEERS 2022 Checklist 29

**Supplemental Methods** **32**

Defining vaccine roll-out rates 32

Further Model Descriptions 33

Fitting Process 34

Characterising behavioural change using data on non-pharmaceutical intervention and mobility 35

Calculating COVID-19 severe, critical, and death cases 36

Lengths of Stay (LoSs) 38

Calculating Disability-adjusted Life Years (DALYs) 40

Adjusting Costs and DALYs over Time 42

Estimating Vaccine Delivery Costs 43

Extrapolating unit costs from base countries to other countries in Africa 52
Extrapolating vaccine unit costs for different roll-out scenarios 54

#### Supplemental Figures

##### Figure S1. Diagram of Transmission Model Structure

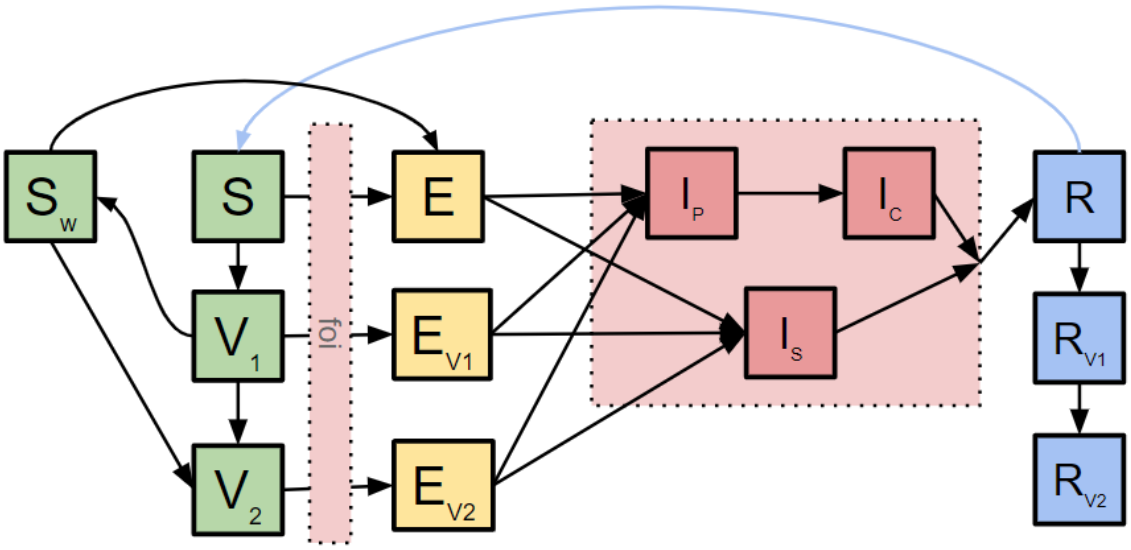


**Caption:** S - Susceptible; V1 - individuals who have received and are protected by one dose of vaccine; Sw - individuals who have received but are not protected by one dose of vaccine; V2 - individuals who have received and are protected by two doses of vaccines; E - exposed individuals; Ev1 - exposed individuals who have previously received and are protected by one dose of vaccines; Ev2 - exposed individuals who have previously received and are protected by two doses of vaccines; Ip - pre-clinical individuals, i.e. individuals who would eventually present symptoms; Ic - clinical and infectious individuals; Is - subclinical (i.e. asymptomatic) and infection individuals; R - removed individuals; Rv1 - individuals with prior infection history and one dose vaccination; Rv2 - individuals with prior infection history and two-dose vaccinations. This model structure and figure have been previously used in Liu et al. (1)

### Figure S2. Performance of model fitting process

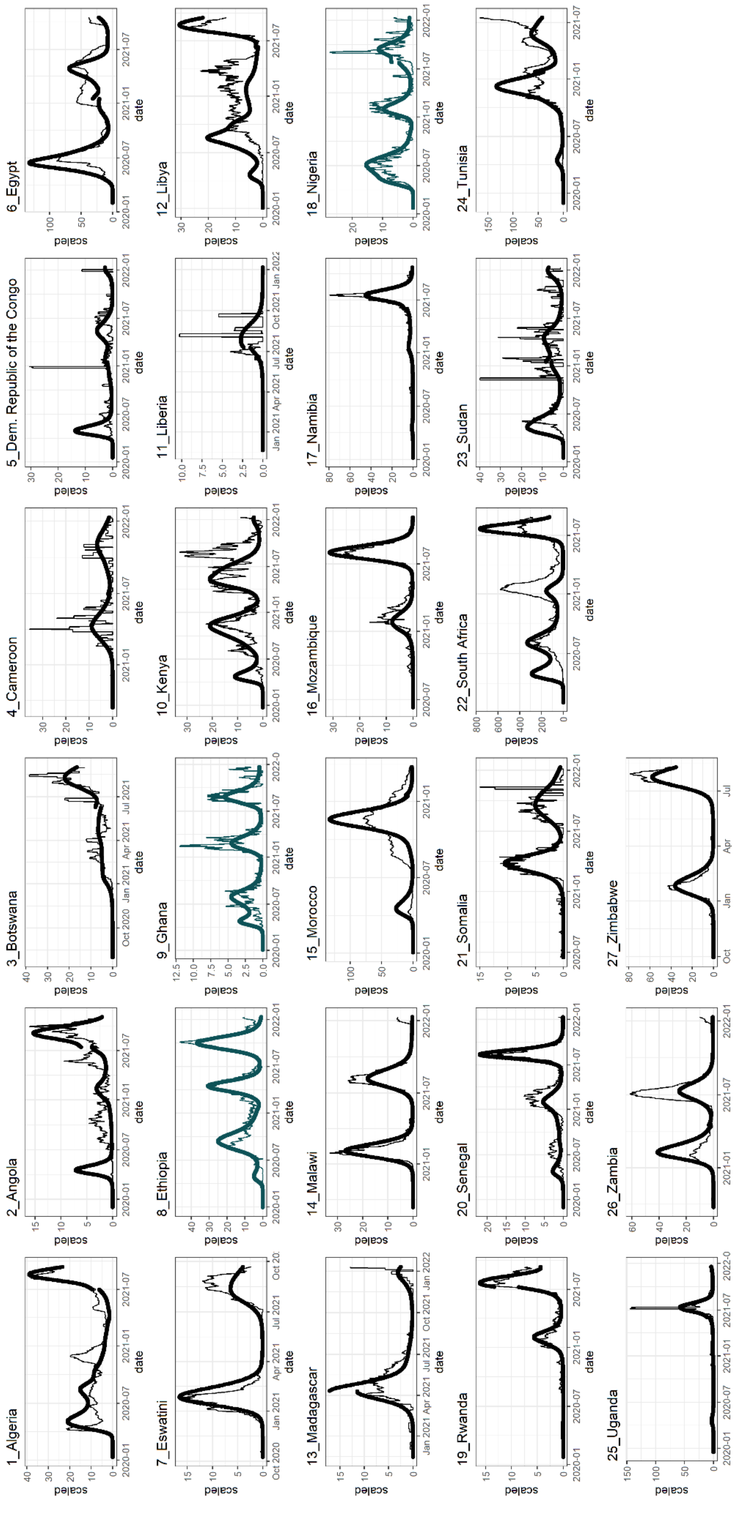


Caption: Countries in dark green indicate countries where we fitted for two VOC introduction dates. All other countries only have one fitted VOC introduction date. The length of the fitting window varies by country and can be found in Supplemental Table S5. The methods involved in generating these results are presented in Supplemental Methods XXXX.

#### Figure S3. Health Outcomes Associated with Different Vaccine Roll-out Scenarios for 27 African Union Members (viral vector vaccines, severe and critical cases)

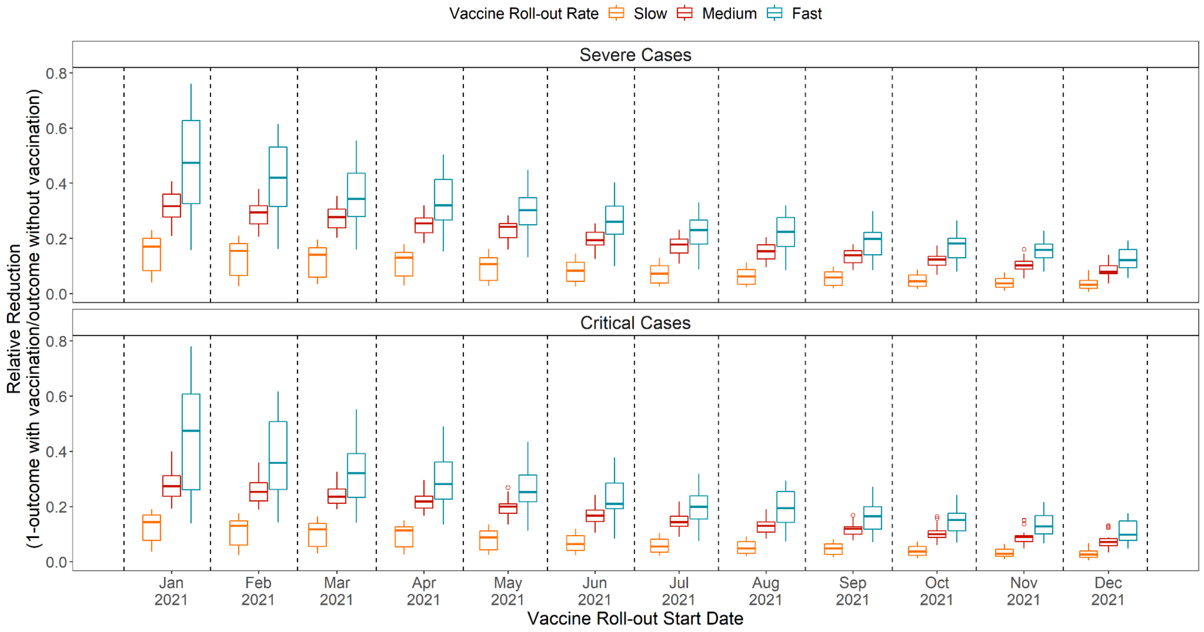


Caption: Relative reduction in health burden as a result of different vaccine roll-out scenarios (i.e. combinations of vaccination program start dates and vaccine roll-out rates) for 27 AU members. Relative reduction is defined as: 1 - outcome with vaccination/outcome without vaccination. Greater relative reductions indicate more effective vaccine roll-out scenarios, and vice versa.

#### Figure S4. Health Outcomes Associated with Different Vaccine Roll-out Scenarios for 27 African Union Members (mRNA vaccines, cases and deaths)

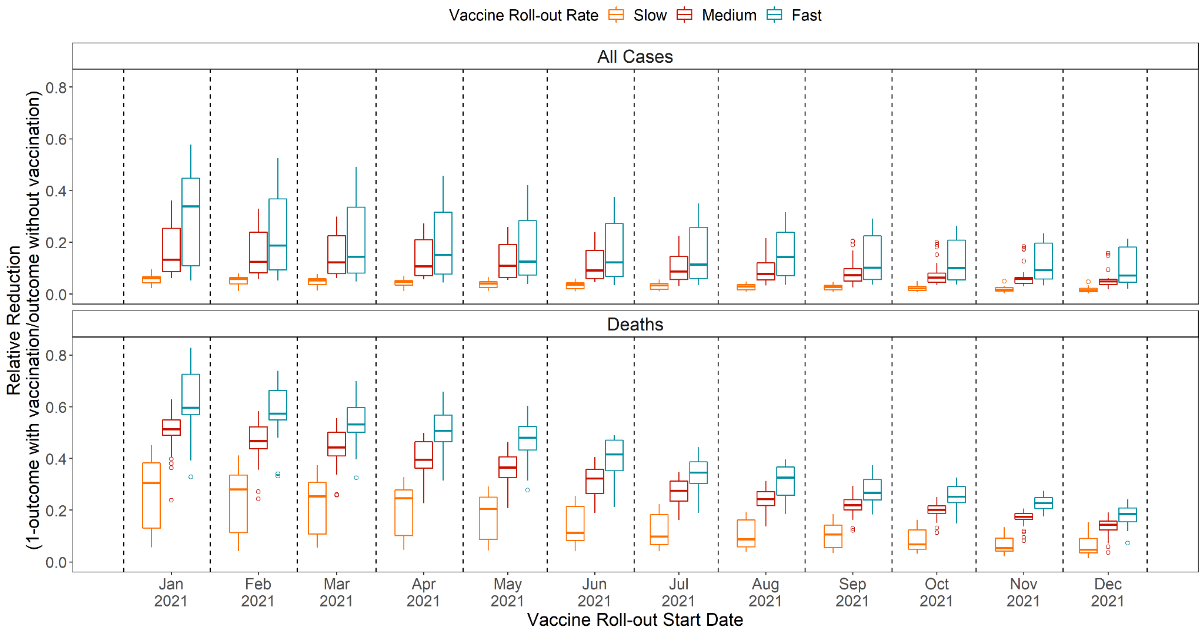


Caption: Relative reduction in health burden as a result of different vaccine roll-out scenarios (i.e. combinations of vaccination program start dates and vaccine roll-out rates) for 27 AU members. Relative reduction is defined as: 1 - outcome with vaccination/outcome without vaccination. Greater relative reductions indicate more effective vaccine roll-out scenarios, and vice versa.

#### Figure S5. Health Outcomes Associated with Different Vaccine Roll-out Scenarios for 27 African Union Members (mRNA vaccines, severe and critical cases)

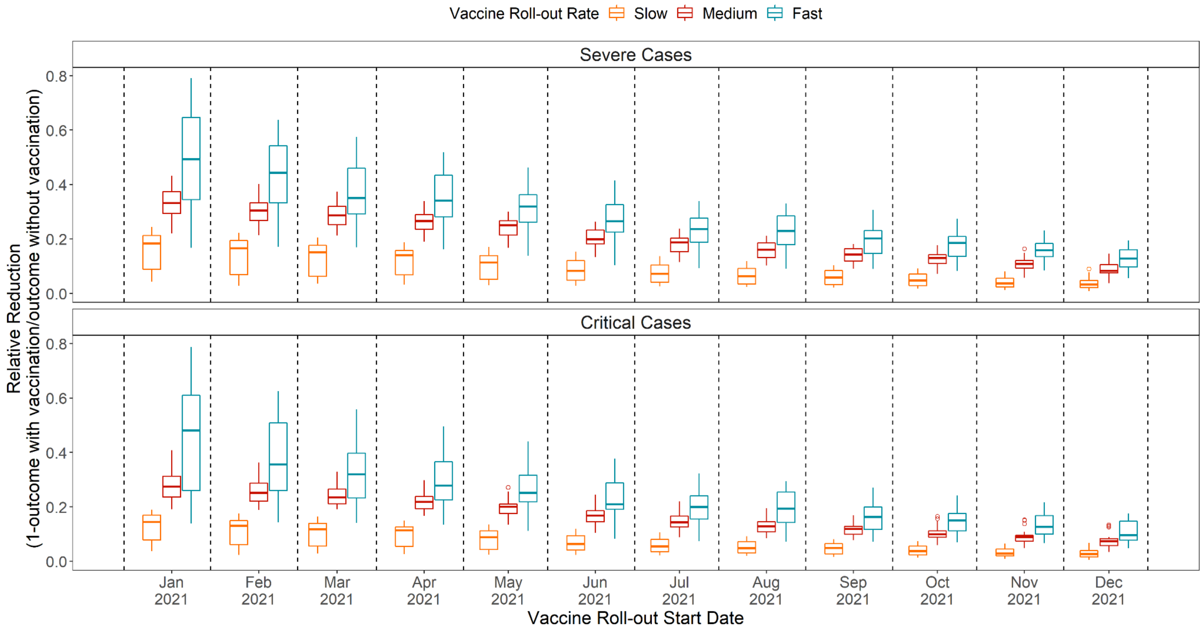


Caption: Relative reduction in health burden as a result of different vaccine roll-out scenarios (i.e. combinations of vaccination program start dates and vaccine roll-out rates) for 27 AU members. Relative reduction is defined as: 1 - outcome with vaccination/outcome without vaccination. Greater relative reductions indicate more effective vaccine roll-out scenarios, and vice versa.

#### Figure S6. Results from sensitivity analyses by vaccine rollout rates: time horizon of outcome aggregation

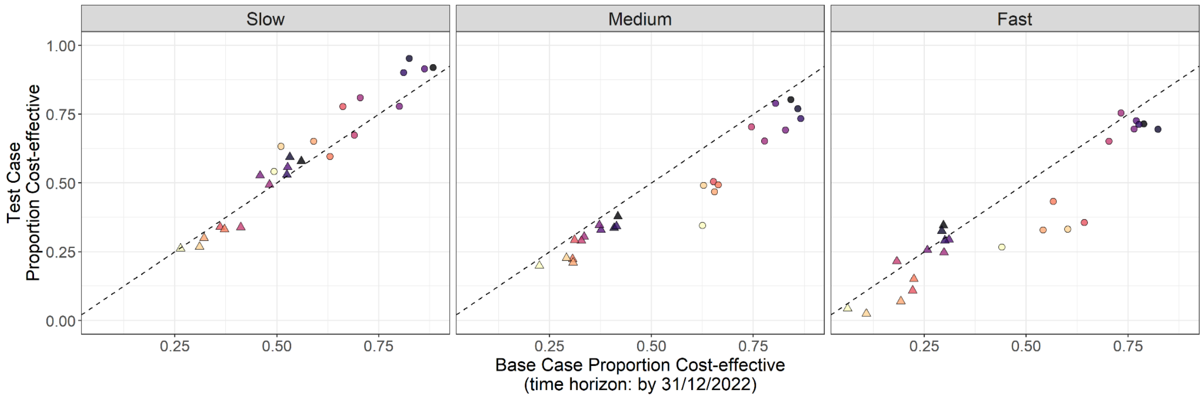


Caption: Point shapes indicate vaccine type, and colours indicate vaccination programme start date. In this figure, each point represents a given vaccine type, vaccination programme start date and vaccine roll-out rate (n = 2*12 points per panel).

#### Figure S7. Results from sensitivity analyses by vaccine rollout rates: vaccine effectiveness estimates

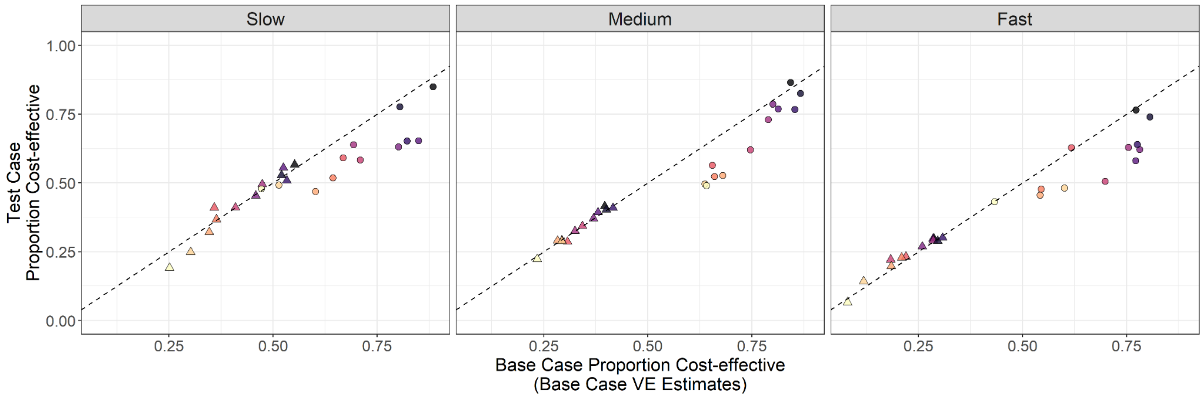


Caption: Point shapes indicate vaccine type, and colours indicate vaccination programme start date. In this figure, each point represents a given vaccine type, vaccination programme start date and vaccine roll-out rate (n = 2*12 points per panel).

#### Figure S8. Results from sensitivity analyses by vaccine rollout rates: willingness-to-pay/ decision-making criteria

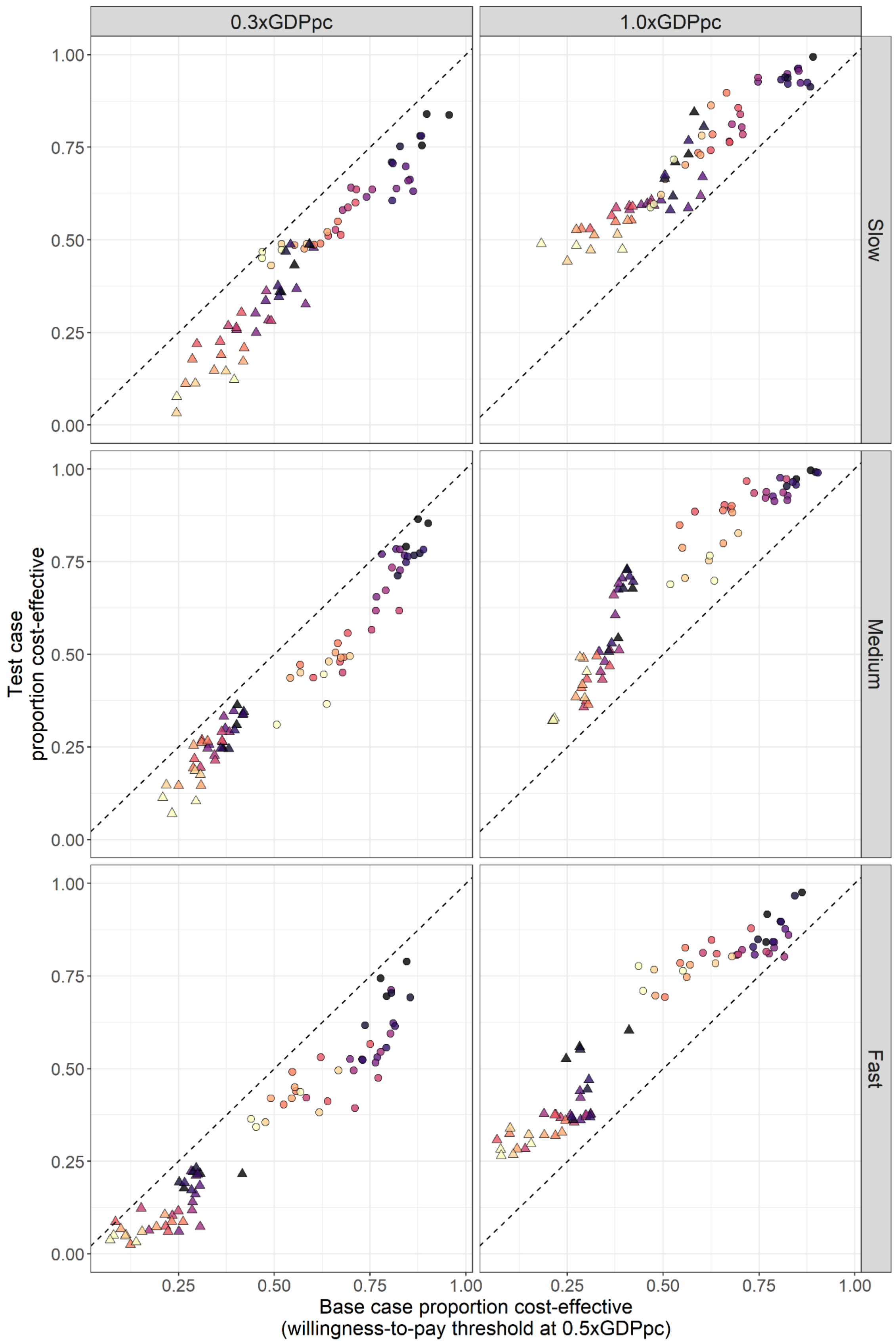

Caption: Point shapes indicate vaccine type, and colours indicate vaccination programme start date. In this figure, each point represents a given vaccine type, vaccination programme start date and vaccine roll-out rate (n = 2*12 points per panel). GDPpc: Gross Domestic Product per capita.

#### Figure S9. Results from sensitivity analyses by vaccine rollout rates: economic evaluation parameter combinations

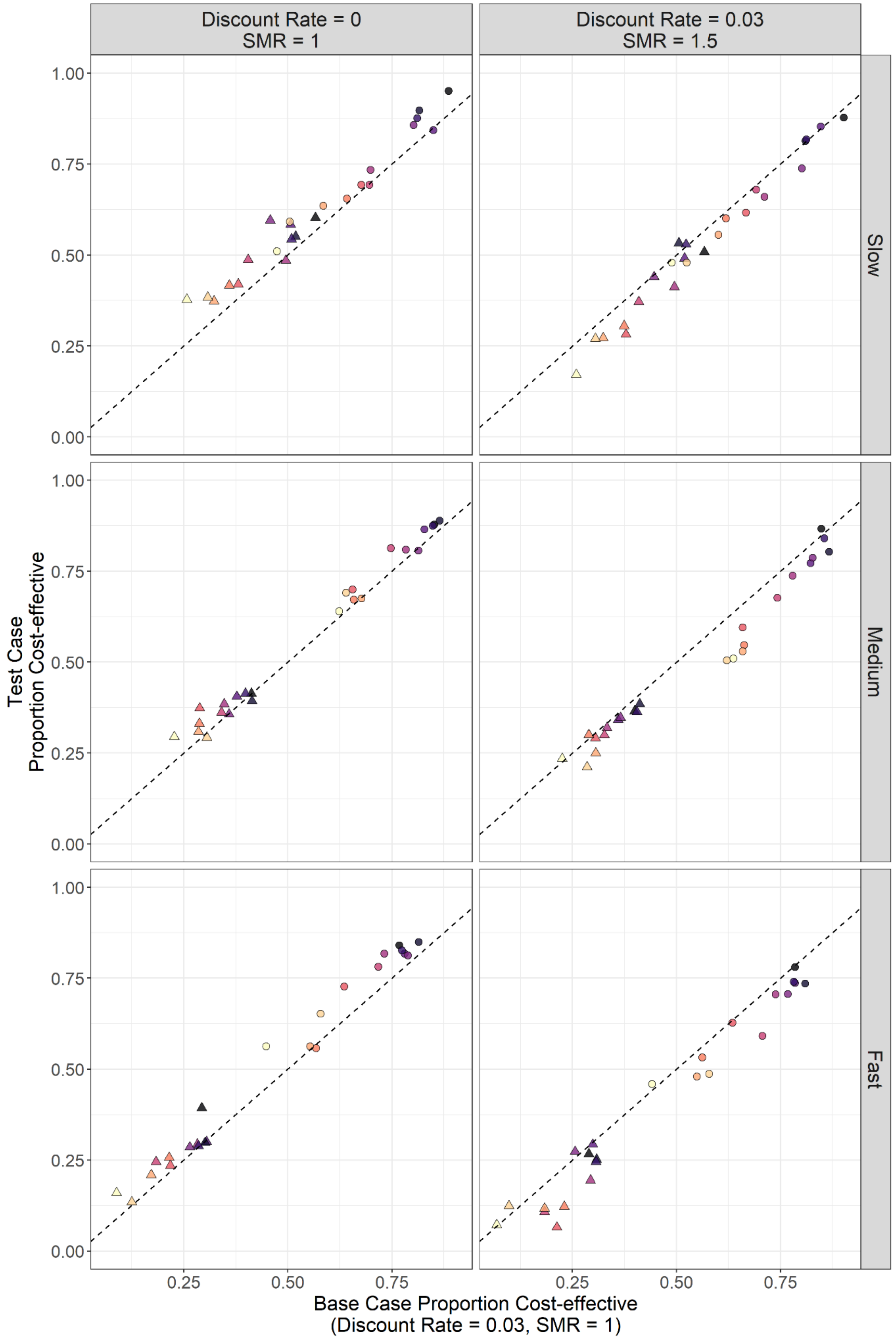

Caption: Point shapes indicate vaccine type, and colours indicate vaccination programme start date. In this figure, each point represents a given vaccine type, vaccination programme start date and vaccine roll-out rate (n = 2*12 points per panel). SMR: standardised mortality ratio.

### Supplemental Tables

#### Table S1. Model Equations

| | | |
|---|---|---|
| $S_i(t)$ | Susceptible individuals among age group $i$ at time $t$ | |
| | $S_i(t+1) = S_i(t)\cdot(1-\lambda_i(t)) - v_{1i}(t+1)\cdot\frac{S_i(t)}{P_{1i}(t)} + R_i(t)\cdot\omega_n$ | [a] |
| $V_{1i}(t)$ | Individuals among age group $i$ who received their first doses at time $t$ | |
| | $V_{1i}(t+1) = V_{1i}(t)\cdot(1-\lambda_i(t)\cdot(1-ve_i) - \omega_v) + v_{1i}(t+1)\cdot\frac{S_i(t)}{P_{1i}(t)} - v_{2i}(t+1)\cdot\frac{V_{1i}(t)}{P_{2i}(t)}$ | [b] |
| $S_{wi}(t)$ | Individuals among age group $i$ who received their first doses but the protection has waned at time $t$ | |
| | $S_{wi}(t+1) = S_{wi}(t)\cdot(1-\lambda_i(t)) + V_{1i}(t)\cdot\omega_v - v_{2i}(t+1)\cdot\frac{S_{wi}(t)}{P_{2i}(t)}$ | [c] |
| $V_{2i}(t)$ | Individuals among age group $i$ who received their second doses at time $t$ | |
| | $V_{2i}(t+1) = V_{2i}(t)\cdot(1-\lambda_i(t)\cdot\left(1-v2e_i\right)) + v_{2i}(t+1)\cdot(\frac{S_{wi}(t)}{P_{2i}(t)} + \frac{V_{1i}(t)}{P_{2i}(t)})$ | [d] |
| $E_i(t)$ | Exposed individuals who are not protected by vaccines (either unvaccinated or have their first doses already waned) among age group $i$ at time $t$ | |
| | $E_i(t+1) = E_i(t)\cdot(1-\sigma) + S_i(t)\cdot\lambda_i(t) + S_{wi}(t)\cdot\lambda_i(t)$ | [e] |
| $E_{vi}(t)$ | Exposed individuals who are protected by one dose of the vaccine among age group $i$ at time $t$ | |
| | $E_{vi}(t+1) = E_{vi}(t)\cdot(1-\sigma) + V_{1i}(t)\cdot\lambda_i(t)\cdot\left(1-ve_i\right)$ | [f] |
| $E_{v2i}(t)$ | Exposed individuals who are protected by one dose of the vaccine among age group $i$ at time $t$ | |

| | | |
|---|---|---|
| | $E_{v2i}(t+1) = E_{v2i}(t) \cdot (1-\sigma) + V_{2i}(t) \cdot \lambda_i(t) \cdot (1 - v2e_i)$ | [g] |
| $I_{pi}(t)$ | Pre-clinical infectious individuals among age group $i$ at time $t$ | |
| | $I_{pi}(t+1) = I_{pi}(t) \cdot (1-\gamma_p) + E_i(t) \cdot \sigma \cdot y_i + E_{vi}(t) \cdot \sigma \cdot y_i \cdot (1 - ve_d) + E_{v2i}(t) \cdot \sigma \cdot y_i \cdot (1 - v2e_d)$ | [h] |
| $I_{ci}(t)$ | Clinical infectious individuals among age group $i$ at time $t$ | |
| | $I_{ci}(t+1) = I_{ci}(t) \cdot \left(1-\gamma_c\right) + I_{pi}(t) \cdot \gamma_p$ | [i] |
| $I_{si}(t)$ | Subclinical infectious individuals among age group $i$ at time $t$ | |
| | $I_{si}(t+1) = I_{si}(t) \cdot (1-\gamma_s) + E_i(t) \cdot \sigma \cdot (1 - y_i) + E_{vi}(t) \cdot \sigma \cdot (1 - y_i \cdot (1 - ve_d)) + E_{v2i}(t) \cdot \sigma \cdot (1 - y_i \cdot (1 - v2e_d))$ | [j] |
| $R_i(t)$ | Recovered individuals | |
| | $R_i(t+1) = R_i(t) \cdot \left(1-\omega_n\right) + I_{si}(t) \cdot \gamma_s + I_{ci}(t) \cdot \gamma_c - v_{1i}(t+1) \cdot \frac{R_i(t)}{P_{1i}}$ | [k] |
| $R_{vi}(t)$ | Individuals who have recovered from their previous infections and have received one dose | |
| | $R_{vi}(t+1) = R_{vi}(t) + v_{1i}(t+1) \cdot \frac{R_i(t)}{P_{1i}} - v_{2i}(t) \cdot \frac{R_{vi}(t)}{P_{2i}(t)}$ | [l] |
| $R_{v2i}(t)$ | Individuals who have recovered from their previous infections and have received two doses | |
| | $R_{v2i}(t+1) = R_{v2i}(t) + v_{2i}(t) \cdot \frac{R_{v2i}(t)}{P_{2i}(t)}$ | [m] |

(continues to the next page)

In which:

| | |
|---|---|
| $\lambda_i(t)$ | Is the force of infection on the population $i$ at time $t$: $$\lambda_i(t) = u_i \cdot \sum_{j=1}^{J} C_{i,j,t} \cdot \frac{(I_{pj}(t) + I_{cj}(t) + I_{sj}(t) \cdot f)}{N_j}$$ Where $j$ depicts age group, $J$ is 16, $f$ is the relative infectiousness of subclinical individuals compared to pre-clinical and clinical individuals (i.e. 50%), and $u_i$ is susceptibility. |
| $P_{1i}(t)$ | Is the population eligible for the first doses in the population $i$ at time $t$: $P_{1i}(t) = N_i - (V_{1i}(t) + S_{wi}(t) + R_{vi}(t) + E_{vi}(t)) - (R_{v2i}(t) + V_{2i}(t)$[illegible] |
| $P_{2i}(t)$ | Is the population eligible for the second doses in the population $i$ at time $t$: $P_{2i}(t) = V_{1i}(t) + S_{wi}(t) + R_{vi}(t) + E_{vi}(t)$ |
| $v_{1i}(t)$ | Is the number of doses to vaccinate the population with dose 1, this is pre-calculated based on vaccine dosing interval strategies. |
| $v_{2i}(t)$ | Is the number of doses to vaccinate the population with dose 2, this is pre-calculated based on vaccine dosing interval strategies. |

**Caption:** This pair of tables has previously appeared in Liu et al. (1)

#### Table S2. Epidemic and Healthcare Process Parameters*

| Parameter | Value | Source |
|---|---|---|
| Age-specific susceptibility ($u_i$) | 0.38 - 0.88 | Davies et al.(2) |
| Age-specific clinical progression rates ($y_i$) | 0.21 - 0.70 | Davies et al(2) |
| Age-specific infection fatality rates | Raw input: 5.2e-6 - 0.13<br>By age group: 6.7e-6 – 8.1e-2 | Levin et al.(3) |
| Age- and country-specific within-population contact pattern ($C_{i,j,0}$) | Country-specific | Prem et al.(4) |
| Country-specific population age structures | Country-specific | United Nations(5) |
| Relationship between mobility and population contact pattern | Defined by linear and nonlinear functions for the *workplace* and *other* settings, respectively. | Davies et al. by fitting to UK data(6) |
| Latent period ($1/\sigma$) | ~gamma (μ = 2.5, k = 2.5) | Pearson et al.(7)<br>Davies et al.(8)<br>Davies et al.(6)<br>Bi et al.(9)<br>Liu et al.(10)<br>Linton et al.(11)<br>Nishiura et al.(12) |
| Duration of preclinical infectiousness ($1/\gamma_p$) | ~gamma (μ = 1.5, k = 4) | |
| Duration of clinical infectiousness ($1/\gamma_c$) | ~gamma (μ = 3.5, k = 4) | |
| Duration of subclinical infectiousness ($1/\gamma_s$) | ~gamma (μ = 5, k = 4) | Assumed, consistent with Davies et al.(8) |
| Relative infectiousness of subclinical infections compared to clinical infections ($f$) | 0.5 | Assumed, consistent with Davies et al.(8) |
| Duration of infection-induced immunity ($1/\omega_n$) | 3 years | Hall et al.(13) |
| Proportion of symptomatic cases that require home-based care | 10% | Torres-Rueda et al. (14) |

* This table is similar to that appeared in the supplemental content of Liu et al. (1)

Table S3. Additional Data Sources*

| Parameter | Source |
| --- | --- |
| Country-level daily COVID-19 Mortality (including 7-day rolling average) | Ritchie et al.(15) |
| Country-level daily COVID-19 vaccine uptake | Ritchie et al.(15) |
| Observed country-specific community mobility | Google(16) |
| COVID-19 Government Response Stringency Index and Government Response Tracker by country | Hale et al.(17) |

* This table has previously appeared in the supplemental content of Liu et al.

Table S4. Other vaccine and vaccination program characteristics

| Parameter | Value | Source |
|---|---|---|
| Supply delay | 4 weeks | Assumed |
| Dosing interval | 4 weeks | WHO (18)<br>UK HSA (19) |
| First dose protection duration ($1/\omega_v$) | 360 days | Assumed |
| Age-specific Prioritisation | 60+ year-olds are prioritised over 20-59. In other words, vaccination among those between 20 and 59 years does not start until the coverage cap among 60+ years has been met. | Liu et al. (20) |
| Maximum willingness to receive vaccines at the population level | 0.7 | WHO (21) |
| Maximum willingness to receive vaccination by age group | 0.6 for those between 20-59 and 0.8 for those above 90 | Assumed based on Robinson et al.(22) |

#### Table S5. Variants of Concern Introduction

| VOC Index | Introduction Date | Single VOC Introduction Countries (n = 24) | | | | | |
|---|---|---|---|---|---|---|---|
| | | Transmissibility | | Severity | | Vaccine Effectiveness/ Efficacy (Against Infection and Disease) | |
| | | Step Change | Overall change | Step Change | Overall change | Step Change | Overall change |
| VOC_1 | - | 1 | 1 | 1 | 1 | 1 | 1 |
| VOC_2 | Fitted | 1.5 | 1.5 | 1.5 | 1.5 | 0.8 | 0.8 |
| VOC_3 | 01/12/2021 | 2.25 | **3.375** | 0.5 | **0.75** | 0.7 | **0.56** |

| VOC Index | Introduction Date | Double VOC Introduction Countries (n = 3) | | | | | |
|---|---|---|---|---|---|---|---|
| | | Transmissibility | | Severity | | Vaccine Effectiveness/ Efficacy (Against Infection and Disease) | |
| | | Step Change | Overall change | Step Change | Overall change | Step Change | Overall change |
| VOC_1 | Fitted | 1.5 | 1.5 | 1 | 1 | 1 | 1 |
| VOC_2 | Fitted | 1.5 | 2.25 | 1.5 | 1.5 | 0.8 | 0.8 |
| VOC_3 | 01/12/2021 | 1.5 | **3.375** | 0.5 | **0.75** | 0.7 | **0.56** |

Caption: VOC_3 is modelled after the Omicron variant. VOC_1 and VOC_2 are pre-Omicron and post-wild type strains. The transmissibility of the pre-Omicron variants are assumed based on evidence in Barnard et al. (23) The transmissibility of the Omicron variant was assumed based on evidence in Pearson et al.(24) The severity of pre-Omicron variants are assumed based on Grint et al. and a Public Health England report. (25,26) The severity of the Omicron variant was assumed based on Danielle Iuliano et al and a UKHSA report. (27,28) The immune-evasiveness of pre-Omicron variants are based on Pouwels et al. and a UK HSA report. (29,30) The immune-evasiveness of the Omicron variant is assumed based on Pearson et al. (24)

Table S6. List of countries with fitted models

| Country Name | ISO3 Code | Fitting Window Starting Date | Fitting Window Ending Date |
|---|---|---|---|
| Algeria | DZA | 25/02/2020 | 12/09/2021 |
| Angola | AGO | 20/03/2020 | 02/11/2021 |
| Botswana | BWA | 30/03/2020 | 01/09/2021 |
| Cameroon | CMR | 06/03/2020 | 11/01/2022 |
| Dem. Republic of the Congo | COD | 11/03/2020 | 11/01/2022 |
| Egypt | EGY | 14/02/2020 | 17/10/2021 |
| Eswatini | SWZ | 14/03/2020 | 22/09/2021 |
| **Ethiopia** | **ETH** | **13/03/2020** | **11/01/2022** |
| **Ghana** | **GHA** | **14/03/2020** | **23/12/2021** |
| Kenya | KEN | 06/03/2020 | 11/01/2022 |
| Liberia | LBR | 17/03/2020 | 07/01/2022 |
| Libya | LBY | 04/03/2020 | 15/09/2021 |
| Madagascar | MDG | 20/03/2020 | 11/01/2022 |
| Malawi | MWI | 29/03/2020 | 11/01/2022 |
| Morocco | MAR | 07/02/2020 | 26/03/2021 |
| Mozambique | MOZ | 22/03/2020 | 04/11/2021 |
| Namibia | NAM | 14/03/2020 | 21/10/2021 |
| **Nigeria** | **NGA** | **28/02/2020** | **11/01/2022** |
| Rwanda | RWA | 14/03/2020 | 10/09/2021 |
| Senegal | SEN | 28/02/2020 | 11/01/2022 |
| Somalia | SOM | 16/03/2020 | 11/01/2022 |
| South Africa | ZAF | 07/02/2020 | 27/08/2021 |
| Sudan | SDN | 13/03/2020 | 11/01/2022 |

| | | | |
|---|---|---|---|
| Tunisia | TUN | 04/03/2020 | 18/07/2021 |
| Uganda | UGA | 21/03/2020 | 20/12/2021 |
| Zambia | ZMB | 18/03/2020 | 11/01/2022 |
| Zimbabwe | ZWE | 20/03/2020 | 10/08/2021 |

Bold font indicates countries with two VOC introduction dates fitted pre-Omicron. All other countries only had one VOC introduction.

#### Table S7. Vaccine Efficacy (Base case)

| Outcome | Vaccine Efficacy | | | |
|---|---|---|---|---|
| | mRNA Vaccines | | Viral Vector Vaccines | |
| | 1st Dose | 2nd Dose | 1st Dose | 2nd Dose |
| Infection | 0.7 | 0.85 | 0.7 | 0.75 |
| Cases[a] | 0.7 | 0.9 | 0.7 | 0.8 |
| Severe cases[b] | 0.85 | 0.95 | 0.85 | 0.9 |
| Critical cases[c] | 0.85 | 0.95 | 0.85 | 0.93 |
| Deaths | 0.85 | 0.95 | 0.85 | 0.95 |
| Onward Transmission | 0.47 | 0.47 | 0.47 | 0.47 |

[a]In the context of this study, cases are defined as symptomatic infections.
[b]In the context of this study, severe cases are defined as those that require hospitalisation.
[c]In the context of this study, critical cases are defined as those that require critical/ intensive cases at some point during their hospital stay. This includes cases that eventually proceed to death as their final outcome.
^

**Caption:** The vaccine efficacy measures used in this study are in line with the measures Barnard et al. (23) However, Barnard et al. (23) did not provide estimates for VE against critical cases. Based on the relative relationship we observed between VE against severe cases (i.e. VE against hospitalisations of at least 2 days long with acute respiratory infection (ARI) code in primary diagnosis field) reported in the UK HSA Vaccine Surveillance Report (Week 12, 24 March 2022), (31) VE against critical cases (i.e. VE against hospitalisations of at least 2 days long with ARI code in primary diagnosis field and with either oxygen, ventilation or ICU use) and VE against deaths, we assumed the VE against critical cases is the average of the VE against severe cases and the VE against deaths.

Table S8. Vaccine Efficacy (lower limit, used for sensitivity analysis)

| Outcome | Vaccine Efficacy | | | |
|---|---|---|---|---|
| | mRNA Vaccines | | Viral Vector Vaccines | |
| | 1st Dose | 2nd Dose | 1st Dose | 2nd Dose |
| Infection | 0.55 | 0.7 | 0.55 | 0.65 |
| Cases[a] | 0.55 | 0.85 | 0.55 | 0.7 |
| Severe cases[b] | 0.75 | 0.9 | 0.75 | 0.8 |
| Critical cases[c] | 0.73 | 0.93 | 0.75 | 0.78 |
| Deaths | 0.7 | 0.95^ | 0.75 | 0.75 |
| Onward Transmission | 0.47^ | 0.47^ | 0.47^ | 0.47^ |

[a]In the context of this study, cases are defined as symptomatic infections.
[b]In the context of this study, severe cases are defined as those that require hospitalisation.
[c]In the context of this study, critical cases are defined as those that require critical/ intensive cases at some point during their hospital stay. This includes cases that eventually proceed to death as their final outcome.
^These values are not varied from their base case values. There is a lack of evidence that supports any variation.

**Caption**: We used this set of alternative VE estimates as a sensitivity analysis. These values were assumed based on a UK HSA report. (32)

Table S9. Itemized cost per dose per activity for base countries - viral vector / AZ-like vaccine - USD$ 2021

| Activity | Ethiopia | Nigeria | South Africa |
|---|---|---|---|
| Planning and coordination | 0.21 | 0.58 | 0.58 |
| Technical Assistance | 0.02 | 0.02 | 0.07 |
| Training | 0.12 | 0.05 | 0.02 |
| Social mobilization | 0.43 | 0.32 | 0.22 |
| Vaccine transport | 0.08 | 0.05 | 0.28 |
| Cold chain | 0.32 | 0.62 | 0.54 |
| PPE | 0.24 | 0.18 | 0.09 |
| Hand hygiene | 0.02 | 0.08 | 0.12 |
| Vaccine dose | 3.93 | 3.93 | 3.93 |
| Vaccine delivery | 0.49 | 0.28 | 0.60 |
| Vaccination certificates | 0.15 | 0.34 | 0.28 |
| Waste management | 0.06 | 0.05 | 0.06 |
| Pharmacovigilance | 0.08 | 0.02 | 0.18 |
| **Unit cost per dose[a]** | **$ 6.16** | **$ 6.52** | **$ 6.96** |

[a] Facility based delivery for 12 months campaign based on country coverage targets

Table S10. Itemized Cost per dose per activity for base countries - mRNA / Pfizer-like vaccine - USD$ 2021

| Activity | Ethiopia | Nigeria | South Africa |
|---|---|---|---|
| Planning and coordination | 0.21 | 0.58 | 0.58 |
| Technical Assistance | 0.02 | 0.02 | 0.07 |
| Training | 0.12 | 0.05 | 0.02 |
| Social mobilization | 0.43 | 0.32 | 0.22 |
| Vaccine transport | 0.03 | 0.05 | 0.28 |
| Cold chain | 0.33 | 0.71 | 0.54 |
| PPE | 0.24 | 0.18 | 0.09 |
| Hand hygiene | 0.02 | 0.08 | 0.12 |
| Vaccine dose | 13.09 | 13.09 | 13.09 |
| Vaccine delivery | 0.49 | 0.28 | 0.60 |
| Vaccination certificates | 0.15 | 0.34 | 0.28 |
| Waste management | 0.06 | 0.05 | 0.06 |
| Pharmacovigilance | 0.08 | 0.02 | 0.18 |
| **Unit cost per dose[a]** | **15.29** | **15.77** | **16.13** |

[a] Facility based delivery for 12 months campaign based on country coverage targets

Table S11. Cost per dose for countries with fitted models - USD$ 2021

| Country Name | ISO3 Code | Viral vector vaccines | mRNA vaccines |
|---|---|---|---|
| Angola | AGO | 7.15 | 16.39 |
| Botswana | BWA | 8.48 | 17.71 |
| Cameroon | CMR | 5.45 | 14.68 |
| Congo - Kinshasa | COD | 6.81 | 16.05 |
| Algeria | DZA | 15.38 | 24.61 |
| Egypt | EGY | 12.68 | 21.91 |
| Ethiopia | ETH | 6.23 | 15.46 |
| Ghana | GHA | 7.02 | 16.25 |
| Kenya | KEN | 11.95 | 21.18 |
| Liberia | LBR | 5.51 | 14.74 |
| Libya | LBY | 14.30 | 23.53 |
| Morocco | MAR | 7.96 | 17.19 |
| Madagascar | MDG | 6.25 | 15.48 |
| Mozambique | MOZ | 5.27 | 14.50 |
| Malawi | MWI | 5.36 | 14.59 |
| Namibia | NAM | 8.37 | 17.60 |
| Nigeria | NGA | 6.90 | 16.13 |
| Rwanda | RWA | 6.05 | 15.28 |
| Sudan | SDN | 6.71 | 15.94 |
| Senegal | SEN | 4.94 | 14.17 |
| Somalia | SOM | 5.38 | 14.61 |
| Eswatini | SWZ | 7.86 | 17.09 |
| Tunisia | TUN | 7.57 | 16.80 |
| Uganda | UGA | 8.75 | 17.98 |
| South Africa | ZAF | 13.83 | 23.06 |
| Zambia | ZMB | 5.99 | 15.22 |
| Zimbabwe | ZWE | 6.85 | 16.08 |

Caption: This is based on facility-based vaccine delivery, medium vaccine roll-out rates and 18-month vaccination campaign duration. These values correspond to the main text Figure 1e.

Table S12. CHEERS 2022 Checklist

| Topic | No. | Item | Location where item is reported |
|---|---|---|---|
| Title | | | |
| | 1 | Identify the study as an economic evaluation and specify the interventions being compared. | **1** |
| Abstract | | | |
| | 2 | Provide a structured summary that highlights context, key methods, results, and alternative analyses. | **2** |
| Introduction | | | |
| Background and objectives | 3 | Give the context for the study, the study question, and its practical relevance for decision making in policy or practice. | **3** |
| Methods | | | |
| Health economic analysis plan | 4 | Indicate whether a health economic analysis plan was developed and where available. | **n/a Not RCT based** |
| Study population | 5 | Describe characteristics of the study population (such as age range, demographics, socioeconomic, or clinical characteristics). | **3-4, Supplemental Table S2-S8** |
| Setting and location | 6 | Provide relevant contextual information that may influence findings. | **4-7** |
| Comparators | 7 | Describe the interventions or strategies being compared and why chosen. | **4-5** |
| Perspective | 8 | State the perspective(s) adopted by the study and why chosen. | **4, 7** |
| Time horizon | 9 | State the time horizon for the study and why appropriate. | **6** |
| Discount rate | 10 | Report the discount rate(s) and reason chosen. | **6-7** |
| Selection of outcomes | 11 | Describe what outcomes were used as the measure(s) of benefit(s) and harm(s). | **5-7** |
| Measurement of outcomes | 12 | Describe how outcomes used to capture benefit(s) and harm(s) were measured. | **6-7** |
| Valuation of outcomes | 13 | Describe the population and methods used to measure and value outcomes. | **6-7** |
| Measurement and valuation of resources and costs | 14 | Describe how costs were valued. | **7, Supplemental Methods p43, p52, p54** |
| Currency, price date, and conversion | 15 | Report the dates of the estimated resource quantities and unit costs, plus the currency and year of conversion. | **7** |
| Rationale and description of model | 16 | If modelling is used, describe in detail and why used. Report if the model is publicly available and where it can be accessed. | **5-6, 8, Supplemental Table S1, Supplemental Figure S1,** |

| | | | |
|---|---|---|---|
| | | | **Supplemental Methods p33-35** |
| Analytics and assumptions | 17 | Describe any methods for analysing or statistically transforming data, any extrapolation methods, and approaches for validating any model used. | **Supplemental Table S11 Supplemental Methods p32, p35, p52, p54** |
| Characterising heterogeneity | 18 | Describe any methods used for estimating how the results of the study vary for subgroups. | **4-6** |
| Characterising distributional effects | 19 | Describe how impacts are distributed across different individuals or adjustments made to reflect priority populations. | **4, Supplemental Methods p33-34** |
| Characterising uncertainty | 20 | Describe methods to characterise any sources of uncertainty in the analysis. | **6-7** |
| Approach to engagement with patients and others affected by the study | 21 | Describe any approaches to engage patients or service recipients, the general public, communities, or stakeholders (such as clinicians or payers) in the design of the study. | **4, 6-7** |
| Results | | | |
| Study parameters | 22 | Report all analytic inputs (such as values, ranges, references) including uncertainty or distributional assumptions. | **4-7 Supplemental Table S1-S11** |
| Summary of main results | 23 | Report the mean values for the main categories of costs and outcomes of interest and summarise them in the most appropriate overall measure. | **8-14** |
| Effect of uncertainty | 24 | Describe how uncertainty about analytic judgments, inputs, or projections affect findings. Report the effect of choice of discount rate and time horizon, if applicable. | **10-14 Supplemental Figures S3-S9** |
| Effect of engagement with patients and others affected by the study | 25 | Report on any difference patient/service recipient, general public, community, or stakeholder involvement made to the approach or findings of the study | **n/a, no patient involved** |
| Discussion | | | |
| Study findings, limitations, generalisability, and current knowledge | 26 | Report key findings, limitations, ethical or equity considerations not captured, and how these could affect patients, policy, or practice. | **14-16** |
| Other relevant information | | | |
| Source of funding | 27 | Describe how the study was funded and any role of the funder in the identification, design, conduct, and reporting of the analysis | **19** |
| Conflicts of interest | 28 | Report authors conflicts of interest according to journal or International Committee of Medical Journal Editors requirements. | **21** |

From: Husereau D, Drummond M, Augustovski F, et al. Consolidated Health Economic Evaluation Reporting Standards 2022 (CHEERS 2022) Explanation and Elaboration: A Report of the ISPOR CHEERS II Good Practices Task Force. Value Health 2022;25. doi:10.1016/j.jval.2021.10.008

### Supplemental Methods

#### Defining vaccine roll-out rates

A simple by-country linear model (cumulative doses per million ~ date) was used to identify the slope of the upward trends observed in cumulative doses per million for each country. Using the coefficients of the `date` variable, we divided the observed vaccine roll-out rate observed in 53 of 55 AU members with relevant data in *Our World in Data* into three categories: slow, medium, and fast. These roll-our rates, by category, have the following summary statistics (doses per million):

| | Min | 1st Q | Median | Mean | 3rd Q | Max |
|---|---|---|---|---|---|---|
| Slow | 8 | 144 | **275** | 246 | 333 | 482 |
| Medium | 499 | 642 | **826** | 817 | 994 | 1268 |
| High | 1359 | 1734 | **2066** | 2685 | 3857 | 5548 |

#### Further Model Descriptions

The conceptual diagram of this model can be found in Supplemental Figure S1. The model input parameters are presented in Supplemental Table S1-4. The adaptation of CovidM that we are using here has been previously described in detail in Liu et al.(1) In short, the model has 13 infection-/ disease-related compartments and age stratification into 16 age groups (ranging from 0-4 to 75+ with five-year increments), totally 208 compartments.

Differences between age groups are characterised by age-specific parameters such as contact rates,(4) susceptibility, clinical fraction,(2) and infection fatality and hospitalisation rates.(3,33)

Differences between countries in the context of the transmission model are characterised by COVID-19 government response stringency index (SI), Google Mobility Index (GM), population age structure, social contact patterns, daily reported COVID-19 deaths. More specifically, SI and GM are used to modify social contact patterns to capture the behavioural changes during the pandemic essential to the transmission of SARS-CoV-2. More details on the method can be found in the Supplemental Methods XXXX.

Compared to a convention SEIR (susceptible-exposed-infectious-removed) model, this adaptation of CovidM:

- can more accurately capture the natural progression of COVID-19 using the "pre-clinical & infectious" and "subclinical & infectious compartments";
- allows both susceptible and removed individuals to receive vaccination. Individuals who received vaccination during an infection is considered "wasted";
- allows for the incorporation of six different types of vaccine effect mechanisms (infection-, disease-, severe disease-, critical disease-, mortality-reducing and onward transmission-preventing);
- allows for the introduction of variants of concerns with changes in susceptibility, severity, and immune evasiveness.
- can more accurately capture the changes in behaviour during the pandemic using SI and GM.

In this model, we assumed that infection-induced immunity among individuals who have never received vaccines and immunity over individuals who had breakthrough infections (vaccinated and then infected) would wane exponentially with an average duration of 3 years. (13) Fully vaccinated individuals without infection history and recovered individuals who then received vaccines do not experience any protection waning effect given this the relatively short time horizon of this study. The lack of waning among recovered individuals who then received vaccines allows us to capture the potential immune enhancement observed in some cases.(34) This setup also improves our capacity in vaccine dose accounting - making sure that most individuals in this model receive only two doses of vaccines.

#### Fitting Process

The model fitting methods used in this study have previously been described in Liu et al. (1) In brief, we estimated the following parameters by fitting this model to country-level daily reported COVID-19 deaths:

1. the basic reproduction numbers (R0). This is relevant for the wildtype strain circulating starting from late 2019/ early 2020.
2. infection introduction dates. Instead of the first day when a case was reported, this is the first day where infection introduction has led to community transmission.
3. COVID-19 death reporting rate. This is the proportion of COVID-19 deaths reported to the surveillance system. If there are 100 COVID-19 deaths but only 10 deaths were reported, the reporting rate is 10%.
4. VOC introduction dates. We only fitted for pre-Omicron VOC introduction dates but not for VOC characteristics. The VOC characteristics we used are based on evidence from the existing literature (see Supplemental Table S5). For 24 countries, one VOC introduction would allow us to capture the epidemic waves. For 3 countries (Ethiopia, Ghana, and Nigeria), two VOC introductions were necessary. The choices between one or two VOC introductions were based on visual examinations of the epidemic peaks captured.

The model was fitted using differential evolution algorithms, implemented using the \`DEoptim\` package in R.(35) We assume daily reported COVID-19 deaths to follow a Poisson distribution.

#### Characterising behavioural change using data on non-pharmaceutical intervention and mobility

In two previous studies, (1,20) we described how we used the Stringency Index (as defined in the context of the Oxford COVID-19 Government Response Tracker) (17) and the Google Mobility Index (16) in great detail. In summary and in the context of this study, for population contact patterns in `work` and `others` settings, we assumed:

1. The relationship between contacts and mobility is defined by functions inferred in Davies et al. based on UK data;(6)
2. The future changes in mobility beyond available data are approximated using a general additive model. Independent variables include day-of-week, country (random effect), an interaction term between day-of-week and mobility type (e.g. grocery, retail, park), Stringency Index (spline), month, and date (spline, to capture slow-varying long-term trend that's not otherwise captured elsewhere).
3. Stringency Index is not expected to vary beyond available data. This is based on the observed Stringency Index in most countries three months prior to the end of the observed time series. The Stringency Index used for future changes range from 19.44 (The Republic of Namibia) to 76.75 (The Republic of Sudan) (high values indicate high stringency).

#### Calculating COVID-19 severe, critical, and death cases

In this study, we projected four health outcomes under different vaccine roll-out scenarios: (1) symptomatic infections; (2) severe cases that require hospitalisation; (3) critical cases that require intensive care unit (ICU) admission; and (4) deaths. The health system parameters that we are using include infection-hospitalisation rate, infection-ICU rate, an infection fatality rate. The values we used for these parameters were estimated in the context of Europe where healthcare resources are considered relatively abundant compared to the rest of the world. For example, the infection-hospitalisation rate is interpreted as the percentage of infections that requires hospital admissions as we assume those who needs hospitalisation were admitted. In the context of the AU, however, the numbers of individuals that requires hospitalisations and those who were admitted are unlikely going to align. Due to scarce health care resources, the cumulative outcomes (2) and (3) are likely going to exceed observation.

Below we provide the procedures that we use to calculate these outcomes. The concepts presented here is similar to our previous studies although we have adapted the equations to capture intermediate outcomes. (1,20)

1. The temporal delays between infection and health outcomes are assumed to follow gamma distributions. The probability density function is capped at 60 days and can be expressed as the following:

   P_severe = *Gamma*(mean = 8.5, shape = 5) + *Gramma*(mean = 14.6, shape = 5)

   P_critical = *Gamma*(mean = 8.5, shape = 5) + *Gramma*(mean = 15.6, shape = 5)

   P_death  = *Gamma*(mean = 26, shape = 5)

   This set of parameters is consistent with that used in Pearson et al. (36)

2. The outcomes that occur on day *t* due to infection on day *t-d* can thus be expressed as:

   Severe(t)_dj = E_j(t-d) * P_severe(d) * IHR +

   Ev1_j(t-d) * P_severe(d) * IHR * (1-VE_severe_1) +

   Ev2_j(t-d) * P_severe(d) * IHR * (1-VE_severe_2)

   Critical(t)_dj = E_j(t-d) * P_critical(d) * IHR +

   Ev1_j(t-d) * P_critical(d) * IHR * (1-VE_critical_1) +

   Ev2_j(t-d) * P_critical(d) * IHR * (1-VE_critical_2)

   Death(t)_dj = E_j(t-d) * P_death(d) * IHR +

Ev1_j(t-d) * P_death(d) * IHR * (1-VE_death_1) +

Ev2_j(t-d) * P_death(d) * IHR * (1-VE_death_2)

where *j* indicates age group.

3. The cumulative outcomes on day *t* can thus be expressed as

   Severe(t) = Sum_{j = 1}{J} * Sum_{d = 1}{60} Severe(t)_dj

   Critical(t) = Sum_{j = 1}{J} * Sum_{d = 1}{60} Critical(t)_dj

   Deaths(t) = Sum_{j = 1}{J} * Sum_{d = 1}{60} Deaths(t)_dj

Note that in this set of formulas we referenced six VE estimates (VE_severe_1, VE_severe_2, VE_critical_1, VE_critical_2, VE_death_1, VE_death_2). They are not values directly from Table S7 and may require further conversion. Only metrics that ends with _o are directly observed. Here, we use the vaccine efficacy against death by the first dose as an example - other VEs (against onon infection outcomes) were converted using the same method.

(1) If m deaths were to occurr without vaccine, in a population vaccinated with the first dose we should avert m*VE_death_1_o deaths.
(2) What has been described in (1) have two components: a. individuals who did not get infected due to vaccination and b. infected individuals whose dead were averted due to vaccination. Component a. can be expressed as m*VE_infection_o. Component b. Can be expressed as m*(1-VE_infection_o)*VE_death_1
(3) With these relationships, we have:

    m*VE_death_1_o = m*VE_infection_o + m*(1-VE_infection_o)*VE_death_1

    With some rearrangement, we have:

    VE_death_1 = (VE_death_1_o - VE_infection_o)/(1-VE_infection_o)

#### Lengths of Stay (LoSs)

We based our parameters on lengths of stay (LoSs) based on Leclerc et al. (37)

| | Beds | n | Stage 1 | Stage 2 | Stage 3 | Mean total lengths of stay (days) |
|---|---|---|---|---|---|---|
| Bed pathways involving CC | CC | 232 | 10.91 | - | - | 10.91 |
| | CC, Ward | 2521 | 13.53 | 7.07 | - | 20.75 |
| | Ward, CC | 183 | 3.39 | 8.77 | - | 12.18 |
| | Ward, CC, Ward | 3603 | 4.1 | 12.25 | 6.9 | 23.32 |
| Bed pathways that do not involve CC | Ward | 29975 | 9.6 | | | 9.60 |

**Table SX.** Lengths of stay by bed pathway in COCIN. (CC = critical care)

We calculated the weighted mean length of stay amongst all bed pathways involving a stay in a critical care bed:

(23.32*3603+12.18*183+20.75*2521+10.91*232)/(232+2521+183+3603) = 21.58 days.

This value was used in calculating the total DALYs (more specifically the YLDs) associated with critical cases. The mean LoS not involving any critical care bed use (i.e. 9.6 days) was used in calculating the total DALYs (more specifically the YLDs) associated with severe cases.

(section continued on the next page)

We calculated the weighted mean ward stay and weighted CC stay among bed pathways involving a stay in a critical care bed:

In ward -
((4.1+6.9)*3603 + 3.39*183 + 7.07*2521 + 0*232)/(232+2521+183+3603) = 8.88 days

In CC -
(12.25*3603 + 8.77*183 + 13.53*2521 + 10.91*232)/(232+2521+183+3603) = 12.60 days

These values, together with the LoSs for bed pathways not involving any stay in a critical care bed (i.e. 9.6 days), were used in calculating the total healthcare costs associated with critical and severe cases, respectively. As a reminder, in the context of this study, severe cases are defined as those that require hospitalisation; critical cases are defined as those that require critical/ intensive care at some point during their hospital stay.

#### Calculating Disability-adjusted Life Years (DALYs)

We calculated DALYs associated with Covid-19 morbidity and mortality as follows:

- We calculated Years Life Lost (YLLs) for Covid-19 deaths using country-specific standard life-tables; (38)
- In the base case we assumed remaining life-expectancy among individuals who die of COVID-19 is the same as the general population of each country, but we varied this in a sensitivity analysis assuming 50% higher baseline mortality rate reflecting the greater prevalence of co-morbidities in people experiencing severe Covid-19; (39)
- For acute morbidity we will use disability weights from for different severity of acute infection and for long-covid using the disability weight for chronic fatigue syndrome (see table below), following as similar approach to Wyper et al. (40)
    - In line with costing assumptions we assume that 10% of community cases seek healthcare and therefore have symptoms corresponding to the moderate disability weight, and that the remaining 90% cases experience mild symptoms for a duration of 5 days.
    - For hospitalised cases we apply the severe weight for 9.60 days for those hospitalised without icu care, and the critical weight for 21.58 days for those hospitalised with a stay in icu (see section above on LOSs)
    - For long-covid we conservatively assume 20% of patients experience post-acute consequences lasting for 6 months.

Disability weighting used for different health states are summarised in the following table adapted from Wyper et al. (40)

| Health state | Description | Disability weight | Source |
|---|---|---|---|
| Asymptomatic | Has infection but does not experience any symptoms | Nil | |
| Mild | Infectious disease, acute, mild: Has a low fever and mild discomfort, but no difficulty with daily activities. | 0.006 (0.002-0.012) | Salomon et al. (41) |
| Moderate (Community; seeking healthcare assistance) | Infectious disease, acute episode, moderate: Has a fever and aches and feels weak, which causes some difficulty with daily activities. | 0.051 (0.032–0.074) | Salomon et al. (41) |
| Severe (Hospitalised; non-intensive care) | Infectious disease, acute episode, severe: Has a high fever and pain, and feels very weak, which causes great difficulty with daily activities. | 0.133 (0.088–0.190) | Salomon et al. (41) |

| | | | |
|---|---|---|---|
| Critical (Hospitalised; intensive care) | Intensive care admission with or without respiratory support. | 0.655 (0.579–0.727) | Haagsma et al. (European Disability Weight Study) (42) |
| Post-acute consequences | Chronic Fatigue Syndrome: Always tired and easily upset. The person feels pain all over the body and is depressed. | 0.219 (0.148–0.308) | Salomon et al. (41) |

#### Adjusting Costs and DALYs over Time

**Adjusting costs to common currency year**

Healthcare costs were estimated in 2019 USD, whereas vaccine delivery costs (including purchase price) were estimated in 2021 USD. To express costs in common currency year we will inflated/deflated care costs and vaccine delivery costs into 2020 USD using the USA GDP deflator:

Cost(target year) = Cost(base year) * deflator(target year)/deflator(base year)

**Discounting**

Costs and DALYs (represented by X in the formula below) occurring in a future year (y) were discounted to the base-line year (y0) of the analysis (2021) using annual discounting:

X(discounted) = X * [1 / ((1 + r)^(y-y0)) ]

where r is the discount rate (0.03 for both costs and DALYs in our base case analysis, and 0 for DALYs in our sensitivity analysis).

#### Estimating Vaccine Delivery Costs

**Estimating unit costs per dose for base countries**

While estimating vaccine unit cost, we covered 12 vaccination activities necessary for the planning, roll-out and delivery of vaccines, as well as the cost of the dose itself. These are planning and coordination, technical assistance, training, social mobilisation, vaccine transport, cold chain, personal protective equipment, hand hygiene, vaccine delivery, vaccination certificates, waste management and pharmacovigilance. The decision to include these 12 activities was based on a model of the costs of delivering the COVID-19 vaccine in the 92 COVAX countries developed by UNICEF.(43) The price per dose per country was obtained from the Graduate Institute Geneva's Covid-19 Vaccine Access that tracks publicly available data on agreements to purchase or supply Covid-19 vaccines.(44)

To estimate the unit cost per dose we used an ingredient-based costing approach.(45) We calculated full economic costs from a health system perspective over several roll-out rates and program duration. We used recent local cost and resource use data from three base countries in Africa: Ethiopia (low-income country (LIC)), Nigeria (lower-middle-income country (lower-MIC)) and South Africa (upper-middle-income country (upper-MIC)). To estimate the resource use and prices of inputs we conducted a literature review, encompassing both peer-reviewed literature and grey literature. An initial description of plausible resource use and list of prices was prepared and shared with public health experts in-country, who reviewed and validated it through several rounds of exercises to arrive at a final description for each sub-activity.

We provide a list of assumptions for the three countries that we collected data for. The author could be contacted for further information about the specific values used.

<table>
<tr><td rowspan="2">Planning and coordination</td><td>Each level of administration will require planning and coordination activities. All planning and coordination staff are assumed to be working with laptops. We assume staff salaries for:<br><br>Ethiopia<ul><li>10 senior government officials at the national level</li><li>10 junior government officials at the national level</li><li>10 senior government officials per region (120 officials in total)</li><li>Two junior government officials per zone (230 officials in total)</li><li>Two junior government officials per woreda (2,108 officials in total)</li><li>One team supervisor per vaccination facility (4,063 supervisors in total)</li></ul></td></tr>
<tr><td>Nigeria<ul><li>20 senior government officials at the national level</li><li>10 senior government officials per state (370 officials in total)</li><li>Seven junior government officials per local area (5,418 officials in total)</li><li>One team supervisor per facility (2,600 supervisors in total)</li></ul></td></tr>
</table>

| | |
|---|---|
| | **South Africa**<br>● 54 senior government officials at the national level<br>● 10 junior government officials at the national level<br>● 25 senior government officials per province (225 officials in total)<br>● Five junior government officials per province (45 officials in total)<br>● 20 government officials per district (1,040 officials in total)<br>● Two junior government officials per local municipality (410 officials in total)<br>● One facility manager per vaccination site (2,335 managers in total) |
| **Technical Assistance** | International consultants provide assistance on all aspects of vaccine rollout, from planning and coordination and social mobilization to cold chain logistics and monitoring and evaluation. We assume all consultants are working with laptops and that teams of consultants require office space in each region. We assume salaries for:<br><br>**Ethiopia**<br>● Eight international consultants for planning and coordination (including assistance with national trainings)<br>● Eight international consultants for monitoring and evaluation<br>● Eight international consultants for service delivery<br>● Eight international consultants for demand generation and communication<br>● Eight international consultants for cold chain & logistics |
| | **Nigeria**<br>● Four consultants for planning and coordination (including assistance with national trainings)<br>● Two consultants for social mobilization<br>● Eight consultants for cold chain and logistics<br>● Five consultants for monitoring and evaluation<br>● Five consultants for service delivery |
| | **South Africa**<br>● Four international consultants for planning and coordination<br>● 17 international consultants for monitoring and evaluation<br>● Five international consultants for service delivery<br>● Four international consultants for demand generation and communication<br>● Two international consultants for cold chain and logistics |
| **Training** | Training is provided in the form of cascade training from national to subnational levels. Assuming bus transport for all participants of trainings, and hall rental, refreshments, and stationery for each training session.We assume staff salaries and per diems for:<br><br>**Ethiopia**<br><br>● One national training of trainers (three days of training): Five senior level government officials and international/local consultants conduct training for teams of junior officials and physicians from each region (five officials per region) |

| | |
|---|---|
| | <ul><li>One training per region (two days of training): conducted by five junior officials per region training three coordinators per zone</li><li>One training per zone (two days of training): conducted by three coordinators per zone training teams of three coordinators per woreda</li><li>One training per woreda (two days of training): conducted by three coordinators per woreda training teams of two coordinators and vaccination team supervisors per facility</li><li>One training per facility (One day of training): conducted by team supervisors training two health extension workers and two nurses per facility</li></ul> |
| | **Nigeria**<ul><li>One national training of trainers (three days of training): Five senior level government officials and four international consultants conduct training for teams of junior level government officials and physicians from 37 states (three junior officials and three physicians per state)</li><li>One training per state (two days of training): conducted by three junior level government officials training teams of three nurses, two vaccinators, and two record keepers for each of the 774 local government areas</li><li>One training per local government area (two days of training): conducted by three nurses training teams of one vaccinator and one record keeper each per health facility</li></ul> |
| | **South Africa**<br>Assuming that all vaccination training is performed online. Individuals involved in vaccination in facility sites must do the training for vaccination sites to be approved. Assume use of laptops for training sessions<ul><li>Training for two nurses (actual average from active vaccination sites), one pharmacist and a data collector per vaccination site (five hours of training)</li><li>Additional vaccination electronic health record (EVDS) training (two hours of training)</li></ul> |
| **Social mobilisation** | **Ethiopia**<ul><li>Two local consultants per region (24 consultants in total) developing the social mobilization strategy and messaging.</li><li>Two support staff and five health extension workers per facility to facilitate community awareness and events.</li><li>National TV ads: 60-second advertisement aired three times daily for 15 days</li><li>National radio ads: 60-second advertisement aired three times daily for 15 days</li><li>Flyers: 1,150,000 printed brochures (10% of coverage target)</li><li>Regional radio ads: 60-second advertisement aired three times daily for 15 days</li><li>Regional TV ads: 60-second advertisement aired three times daily for 15 days</li></ul> |

| | |
|---|---|
| | • 48 audio-mounted vehicles used for 15 days<br>• Banners and posters at each vaccination site<br>• Advocacy workshops and capacity building workshops conducted at national, regional, zone, and woreda levels |
| | **Nigeria**<br>• One local consultant per state developing the social mobilization strategy and messaging.<br>• One local worker and one local leader per health facility to facilitate community awareness and events.<br>• National TV ads: 60 second advertisement aired daily for six months<br>• National radio ads: 60 second advertisement aired daily for six months<br>• Flyers: 200 printed flyers per health facility<br>• Local radio ads (at each local area): 60 second advertisement aired daily for one month |
| | **South Africa**<br>• 41 members of a ministerial advisory committee on social change, aimed at driving social behaviour change in communities, providing support<br>• 14 members of the Government Communication Information System (GCIS) lead the communication response.<br>• Two GCIS Provincial Directors provide regional support<br>• One councillor per local municipality and two GCIS workers per ward to facilitate community awareness and events<br>• TV and radio ads at national and provincial level<br>• Flyers and posters at facility level |

| | |
|---|---|
| **Vaccine transport** | We assume full cold storage capacity at the national level for a single shipment of all doses, and we have allocated numbers of doses equally across regions. Storage assumptions:<br><br>● Volume per vaccine dose : 3.76cm^3<br>● Refrigerated truck volume: 30,000L<br>● Hilux double cab pickup volume (truck bed): 1,204L<br>● Refrigerated truck vaccine storage capacity: 1,563 boxes per truck, 3,751,200 doses<br>● Hilux truck vaccine storage capacity: five boxes per truck, 23,920 doses<br>● Vaccine carrier volume (from woreda to facility): 2.7L, 717 doses<br>● Fuel efficiency: 10km/L<br>● Vaccines are picked up from local area refrigerators by motorcycle and delivered to facilities using long-range vaccine carriers (2.7L).<br><br>**Ethiopia**<br><br>● Five refrigerated trucks (and five drivers) at the national level<br>● 1,493 hilux double cab trucks (and 1,493 drivers) at the zone and woreda levels<br>● Average distance from regional capital to country capital: 554.2km<br>● Average distance from zone capital to regional capital: 315.97km<br>● Average distance from woreda to zone capital: 353.86km<br>● Average distance from health facility to woreda: 34.3km<br>● Average catchment area of each health facility: 278.7km2<br>● Number of deliveries in country (to national stores): nine<br>● Number of deliveries per region by refrigerated truck: five (assuming four days for roundtrip delivery)<br>● Number of deliveries per zone by hilux truck: 26 (assuming two days for roundtrip delivery)<br>● Number of deliveries per woreda by hilux truck: 21 (assuming two days for roundtrip delivery)<br>● Number of deliveries per facility by motorcycle: 33 (assuming one day for roundtrip delivery) |
| | **Nigeria**<br>● Three refrigerated trucks (and three drivers) at the national level<br>● 1,659 hilux double cab trucks (and 1,659 drivers) at the local area and ward level<br>● Average distance from state capital to Abuja: 515km<br>● Average distance from local area capital to state capital: 109km<br>● Average distance from health facility (or ward) to local area capital: 19km<br>● Number of deliveries in country (to national stores): three<br>● Number of deliveries per state by refrigerated truck: three (assuming four days for delivery)<br>● Number of deliveries per local area by truck: seven (assuming two days for delivery) |

| | |
|---|---|
| | ● Number of deliveries per ward by truck: three (assuming two days for delivery)<br>● Number of deliveries per facility by motorcycle: 98 (assuming one day for delivery) |
| | **South Africa**<br>● Two drivers with two refrigerated trucks take the vaccine doses from OR Tambo Airport to Biovac National Warehouse<br>● Distance from the airport to the warehouse is 30 km.<br>● 17 drivers with 17 refrigerated trucks take the vaccine doses from Biovac National Warehouse to provincial distribution depots.<br>● 2,336 drivers with hilux trucks deliver the doses from the provincial warehouses to each vaccination site (2336 vaccination sites in total). Vaccines are transported in large long range cold boxes<br>● Each driver does 14 deliveries for single dose vaccines and 21 for double dose vaccines.<br>● One day for delivery to national warehouse and facilities, four days (return) for delivery from national warehouse to provincial depots<br>● Average distance from Biovac national warehouse to provincial depots: 714 km<br>● Average distance from provincial depots to health facility: 129 km |
| **Cold chain** | We assume office space rented at national and subnational level<br><br>**Ethiopia**<br>● 6.5 10,000L cold rooms at the national level<br>● One 10,000L cold room per region (12 in total)<br>● One 92L solar direct drive fridge per zone<br>● One 50L solar direct drive fridge in one quarter of woredas<br>● Each main power refrigerator uses 19,272 kilowatt-hours per year<br>● Each solar direct drive refrigerator uses 12,176.4 kilowatt-hours per year<br>● Each cold room uses 475,668 kilowatt-hours per year<br><br>Cold chain and logistics staff:<br>● 10 senior-level government officials at national level<br>● 10 junior-level government officials at national level<br>● 10 senior-level government officials per region (120 officials in total)<br>● 10 junior-level government officials per region (120 officials in total)<br>● Three junior-level government officials per zone (345 officials in total)<br>● One junior-level government officials per woreda (1054 officials in total) |

| | |
|---|---|
|  | **Nigeria**<br>● 15 30,000L cold rooms at the national level<br>● One 10,000L cold room per state (37 in total)<br>● One 145L main power fridges per state (37 in total)<br>● One 92L solar direct drive fridge per local area (774 in total)<br>● 0.5 50L solar direct drive fridge per ward (50% of wards have fridge - 4783 in total)<br>● Each main power refrigerator uses 19,272 kilowatt-hours per year<br>● Each solar direct drive refrigerator uses 12,176.4 kilowatt-hours per year<br>● Each cold room uses 475,668 kilowatt-hours per year<br><br>Cold chain and logistics staff:<br>● National logistics working group: six members at national level and 37 at state level<br>● State logistics working group: 10 members per state (370 in total)<br>● Local area logistics working group: three members per local area (2322 in total)<br>● Cold chain teams composed of one cold chain officer, one logistics officer, and one immunization officer for six zones, 37 state, and 774 local government areas |
|  | **South Africa**<br>● Three cold rooms (10,000L) at the national level are devoted to COVID-19 vaccine doses at BIOVAC National Warehouse<br>● 18 cold rooms (10,000L) at the provincial level.<br>● Eight ultra-cold storage freezers (528L) for ultra-cold storage vaccines at Biovac National Warehouse<br>● One ultra-cold storage freezer per province<br>● One Solar direct drive Meta Fridge, 50L per vaccination site (2336)<br>● Each main power refrigerator uses 19,272 kilowatt-hours per year<br>● Each solar direct drive refrigerator uses 12,176.4 kilowatt-hours per year<br>● Each cold room uses 475,668 kilowatt-hours per year<br><br>Cold chain and logistics staff:<br>● 30 cold chain professionals at the national level<br>● 15 cold chain professionals at provincial depots (255 in total)<br>● One pharmacist at facility level |
| **PPE** | Assuming only surgical masks and examination gloves used: three surgical masks per person per day and 10 pairs of gloves per vaccinator per day. |
| **Hand hygiene** | Hands washed before each dose: we assume 50% of handwashing is done with soap and water, and 50% of handwashing done with hand sanitiser.<br>Facilities use water taps<br>1 litre of water used per hand wash, 1 mL of soap per hand wash. 3 ml of hand sanitiser per hand wash. |
| **Vaccine dose** | Assumptions: |

| | |
|---|---|
| | ● 15% wastage<br>● 10% markup for freight cost (cost of delivering doses to country) |
| **Vaccine delivery** | **Ethiopia**<br>● 14 days of vaccine delivery annually, split into two 7-day campaigns<br>● Doses delivered in facilities by two nurses, one health extension worker, one data collector, and one support staff per site per day<br>● One syringe, one alcohol swab, plaster, and dry swab per dose<br>● 200 doses delivered per facility per day (1 minute spent handwashing, 5 minutes per dose)<br>● Two tables in facility-based delivery, six chairs staff members plus 10 chairs for waiting area and for individual receiving vaccine |
| | **Nigeria**<br>● Doses delivered in facilities by three nurses, two record keepers per site per day<br>● One syringe, one alcohol swab, plaster, and dry swab per dose<br>● 1 minute spent handwashing, 5 minutes per dose<br>● Two tables in facility-based delivery, five chairs for staff members plus 10 chairs for waiting area and for individual receiving vaccine |
| | **South Africa**<br>● Doses delivered in facilities by two nurses per site per day<br>● One syringe, one alcohol swab, plaster, and dry swab per dose<br>● 100 doses delivered per facility per day (1 minute spent handwashing, 5 minutes per dose)<br>● Two tables in facility-based delivery, chairs for each staff member plus 10 for waiting area and for individual receiving vaccine |
| **Vaccination certificates** | ● One certificate per vaccinated individual, assuming 3 minutes for record keeping per dose.<br>● One FTE staff member per local area level entering data with a laptop into national vaccine database and office space |
| **Waste management** | ● 5L safety box/sharps container: can contain 100 0.5ml syringes (20 syringes per nominal litre).<br>● One biohazardous bag per delivery site per day for used PPE |
| **Pharmacovigilance** | Assuming all staff are working with laptops, and with office space per zone and per region. Assuming a rate of 12.98 serious adverse events per 100,000 doses, each serious AEFI would require 30 minutes of nurse time for management. We assume 15 chairs per vaccination site for vaccinated individuals to wait for 15 mins following immunization.<br><br>**Ethiopia**<br>● One record keeper for data entry, monitoring, and evaluation per zone (73 in total)<br>● One data manager per region (12 in total) overseeing zone record keepers. |

| | |
|---|---|
| | <ul><li>One AEFI (adverse event following immunization) kit per month</li><li>One AEFI reporting form and one case investigation form per AEFI</li></ul> |
| | **Nigeria**<ul><li>One record keeper for data entry, monitoring, and evaluation per state (37 in total)</li><li>One data manager overseeing 37 record keepers.</li><li>One AEFI kit per month</li></ul> |
| | **South Africa**<br>One data collector for data entry, monitoring and evaluation per local municipality (205 in total)<br>One data manager per province overseeing local data collectors.<br>One AEFI kit per month<br>One AEFI reporting form and one case investigation form per AEFI |

#### Extrapolating unit costs from base countries to other countries in Africa

To calculate the costs for other African countries, we extrapolated our unit cost per dose estimates for Ethiopia, Nigeria and South Africa to LICs, lower-MICs and upper-MICs, respectively, based on country-specific resource use and health systems data and standard approaches to adjusting prices. Each cost input in the ingredients costing was classified as a tradeable good, non-tradeable good or staff cost.(46)

Tradeable goods are generally defined as those that can easily be traded in the international market and include goods such as medical or other supplies and medications. To convert costs of tradeable goods from the base country (eg, South Africa) to a 'second' country (eg, Namibia), we first converted the prices from local currency to 2021 US$ and then apportioned the percentage of the unit cost that was composed of tradeable goods in 2021 US$ from the base country to the second country.

Non-tradeable goods cannot be easily traded in international markets and generally need to be consumed in the country where they have been produced (eg, buildings and utilities). To convert these, we multiplied the proportion of the unit cost that was defined as non-tradeable (in 2021 US$) by the ratio between the 2021 GDP per capita (adjusted for purchasing power parity, or 'PPP') of the second country and the 2021 GDP per capita (adjusted for PPP) of the base country. Data on GDP per capita (adjusted for PPP) were found in the World Bank database.(47)

To convert staff costs from a base country to a second country, we used conversion rates from a regression analysis on wages of health workers for 193 countries to predict wages by country income category relative to GDP per capita.(48) Country-specific wages for physicians, nurses and other health workers were estimated using the conversion rates and respective GDP per capita. Staff costs were extrapolated by multiplying the staff proportion of the unit cost by the ratio between the estimated wages and actual salary levels from the base countries.

Unit costs used for other countries were obtained through extrapolation of these three countries based on country income groups (i.e. LIC, LMIC or UMIC) and cost types (i.e. costs on tradable goods, non-tradable goods, or staff). Tradeable costs were assumed to be constant across countries (e.g. vaccine prices), with non-tradable costs adjusted by country income level. This approach assumes that resource use would be the same within country income categories (adjusted by population and size) but prices of inputs would vary by country. (46)

We further adjusted the unit cost per dose to account for the feasible scale of the vaccine roll-out and delivery in each country. Seven activities were adjusted according to scale per country including planning and coordination, technical assistance, training, social mobilisation, vaccine transport, cold chain and pharmacovigilance.

Each cost input for the base countries was assigned a sub-activity level according to where COVID-19 vaccine-specific resources are deployed - first level (eg, national), second level (eg, province), third level (eg, district) and facility level.  The type and number of administrative divisions for countries in Africa were gathered from a variety of government and non-government sources, and the number of public health facilities per country was obtained from a comprehensive spatial inventory of public health facilities for 50 countries in Sub-Saharan Africa. (49)

The type of administrative division in the other countries was matched to their respective base country's sub-activity level and the unit cost per dose apportioned accordingly. The unit cost at the sub-activity level was multiplied by an adjustment factor capturing the difference in the amount of administrative divisions and health facilities between each country and the base country.

#### Extrapolating vaccine unit costs for different roll-out scenarios

From the cross-country vaccine unit costs cross-extrapolation step, we obtained country level vaccine unit costs by three different roll-out rate levels (275, 826, 2066 doses/ million population-day), two vaccine types (viral vector and mRNA vaccines), and various different program duration length (1 year, 1.5 years, 2 years, 2.5 years). With this raw data we derived a linear model with country-specific coefficient:

(vaccine unit cost) = a + b1*ISO3C + b2*VaccineType +
b3*ProgrammeDuration + b4*RolloutRate

ISO3c and VaccineType are categorical variables; ProgrammeDuration and Rollout Rate are continuous variables. We found that all variable returns statistical significance. This model has a multiple R-squared of 0.9324 and adjusted R-squared of 0.9277. We used this model to extrapolate vaccine unit costs for additional roll-out scenarios.

Moreover, based on the results from the regression model, we learned that mRNA is positively associated with vaccine unit cost. Both ProgrammeDuration and RolloutRate are negatively associated with the vaccine unit cost due to large initial fixed costs and smaller variable costs.

26. Public Health England. SARS-CoV-2 variants of concern and variants under investigation in England - Technical Briefing 14. Public Health England; 2021 Jun.

27. UK Health Security Agency. SARS-CoV-2 variants of concern and variants under investigation in England - Technical briefing: Update on hospitalisation and vaccine effectiveness for Omicron VOC-21NOV-01 (B.1.1.529) . UK Health Security Agency; 2021 Dec.

28. Iuliano AD, Brunkard JM, Boehmer TK, Peterson E, Adjei S, Binder AM, et al. Trends in Disease Severity and Health Care Utilization During the Early Omicron Variant Period Compared with Previous SARS-CoV-2 High Transmission Periods - United States, December 2020-January 2022. MMWR Morb Mortal Wkly Rep. 2022 Jan 28;71(4):146–52.

29. Pouwels KB, Pritchard E, Matthews PC, Stoesser N, Eyre DW, Vihta K-D, et al. Effect of Delta variant on viral burden and vaccine effectiveness against new SARS-CoV-2 infections in the UK. Nat Med. 2021 Dec;27(12):2127–35.

30. UK Health Security Agency. COVID-19 vaccine surveillance report Week 2. UK Health Security Agency; 2022 Jan.

31. UK HSA -. COVID - 19 vaccine surveillance report (Week 12) [Internet]. UK HSA; 2022 Mar [cited 2022 Apr 7]. Available from: https://sciwheel.com/work/item/12766743/resources/14134508/pdf

32. Public Health England. COVID-19 vaccine surveillance report Week 36. Public Health England; 2021 Sep.

40. Wyper GMA, Assunção RMA, Colzani E, Grant I, Haagsma JA, Lagerweij G, et al. Burden of Disease Methods: A Guide to Calculate COVID-19 Disability-Adjusted Life

Years. Int J Public Health. 2021 Mar 5;66:619011.
